# Efficacy of Personalized Connectivity-Guided Accelerated Brain Stimulation in a Naturalistic Treatment-Resistant Depression Population

**DOI:** 10.64898/2026.01.11.26343900

**Authors:** Ru Kong, Xiao Wei Tan, Aihuiping Xue, Leon Qi Rong Ooi, Shih Ee Goh, Jonathan Jie Lee, Jingwen Cheng, Trevor Wei Kiat Tan, Rachel Si Yun Tan, Jovi Zheng Jie Koh, Hasvinjit Kaur Gulwant Singh, Jia Hui Chai, Luming Shi, Shan Siddiqi, B.T. Thomas Yeo, Phern-Chern Tor

## Abstract

Personalized connectivity-guided accelerated intermittent theta burst stimulation (iTBS) has shown promising efficacy for treatment-resistant depression (TRD) in carefully selected trial populations. Evidence remains limited in naturalistic TRD patients with complex psychiatric comorbidities often excluded from such trials. Here, we report an open-label, single-arm proof-of-concept study of a personalized connectivity-guided targeting approach, TAO-TMS (Tree-based Algorithm for Optimized Transcranial Magnetic Stimulation), in a naturalistic Asian TRD sample. Participants averaged 1.7 psychiatric comorbidities, including personality disorders, autism and obsessive-compulsive disorder. Of 21 enrolled participants, 20 completed 50 iTBS sessions over 5 days. The primary outcome was response rate (≥50% reduction in Montgomery-Åsberg Depression Rating Scale score) immediately after treatment. Among participants with available data, response rates were 65% immediately after treatment, 50% at 1 month and 31% at 3 months. In exploratory analyses, 70% responded within 4 weeks post-treatment; the corresponding rate was 83% in a subgroup of participants meeting typical randomized-trial eligibility criteria (N = 12). For descriptive local context, we note that a retrospective cohort at the same hospital receiving non-accelerated TMS with scalp-based BeamF3 targeting had an immediate response rate of 22%. Post hoc electric-field modeling confirmed that TAO-TMS achieved the intended targeting of attention networks. Statistically significant functional connectivity changes were observed in each participant, but varied widely across participants. Exploratory economic modeling suggested lower one-year costs for TAO-TMS than electroconvulsive therapy, with similar quality-adjusted life-years. TAO-TMS was feasible and well tolerated. Its preliminary clinical signal warrants evaluation in randomized controlled trials.

## Introduction

Treatment-resistant depression (TRD) is defined as major depressive disorder (MDD) that fails to respond to adequate trials of at least two antidepressant medications (Thase and Rush, 1997; Conway et al., 2017; Tor et al., 2022). 30% of patients with MDD are estimated to be treatment-resistant (Rush et al., 2006; McIntyre et al., 2023). TRD represents a significant public health burden due to its profound impact on quality of life and high healthcare utilization (Ho et al., 2013; Johnston et al., 2019; Chodavadia et al., 2023). Conventional pharmacological and psychotherapeutic approaches often prove insufficient for TRD, necessitating neurostimulation techniques, such as transcranial magnetic stimulation (TMS; George et al., 2013) or electroconvulsive therapy (ECT; Sackeim et al., 2001). TMS operates by inducing localized electric currents in brain tissues, modulating cortical excitability (Huerta and Volpe, 2009; Fox et al., 2012). Accelerated intermittent theta burst stimulation (iTBS), a more efficient variant of repetitive TMS, condenses sessions into shorter treatment windows, accelerating treatment timelines (Huang and Rothwell, 2004; Blumberger et al., 2018; Cole et al., 2020; Chu et al., 2021).

The therapeutic potential of TMS has been further advanced through individualized connectivity-guided targeting approaches (Fox et al., 2012; Liston et al., 2014; Bergmann et al., 2016; Cash et al., 2021; Siddiqi et al., 2021; Lynch et al., 2022; Hearne et al., 2025). For example, the Stanford Accelerated Intelligent Neuromodulation Therapy (SNT) integrates personalized targets with a 5-day accelerated iTBS protocol, yielding exceptionally high response rates in TRD (Cole et al., 2020, 2022). Subsequent trials have suggested that personalized TMS targets may outperform scalp-based targets in TRD (Li et al., 2026; Taylor et al., 2026a). However, previous TRD trials have generally used restrictive eligibility criteria that excluded several psychiatric comorbidities commonly encountered in clinical practice (Cole et al., 2022; Li et al., 2024, 2026; Taylor et al., 2026a). Consequently, the feasibility and clinical outcomes of accelerated, connectivity-guided TMS remain underexplored in naturalistic TRD populations.

To address these gaps, we applied our previously developed personalized targeting procedure, TAO-TMS (Tree-based Algorithm for Optimized Transcranial Magnetic Stimulation; Kong et al., 2026), in an open-label trial of naturalistic treatment-resistant depression (TRD) patients in Singapore (NCT06341803). In our previous study, TAO-TMS targets compared favorably with other personalized targets (Fox et al., 2013; Cash et al., 2021) in terms of test-retest reliability, scalp proximity and anticorrelations with the subgenual anterior cingulate cortex (Kong et al., 2026). Because TAO-TMS identifies near-scalp targets, the present protocol was designed to use lower stimulation intensities (Stokes et al., 2005), with the aim of reducing scalp discomfort. The patient population examined here is clinically complex, with a higher psychiatric comorbidity burden than populations enrolled in previous accelerated connectivity-guided iTBS trials, as well as substantial polypharmacy, and variable episode durations. Notably, a retrospective study from the same hospital reported a 22% response rate immediately after non-accelerated BeamF3 TMS (Ye et al., 2024), suggesting that the present sample represents a population with lower historical TMS responsiveness than those commonly described in published studies.

As in the SNT protocol, each participant received 50 iTBS sessions (1,800 pulses; 10 sessions/day over 5 days) delivered using a neuronavigated TMS system. As in SNT, stimulation intensity was linearly adjusted for the cortical depth of the target relative to the motor hotspot (Stokes et al., 2005). We additionally used post hoc electric field modeling to characterize the network composition and baseline sACC connectivity of TAO-TMS hotspots, using BeamF3 as a conventional anatomical reference. We also assessed pre-treatment to post-treatment functional connectivity (FC) changes, and performed an exploratory health economic comparison with ECT. This proof-of-concept study evaluated the feasibility, tolerability as measured by treatment completion, preliminary clinical outcomes, and targeting characteristics of TAO-TMS in a naturalistic TRD population with complex psychiatric comorbidities.

## Methods and Materials

### Participants and procedure

A preregistered open-label trial was conducted at the Institute of Mental Health (IMH), Singapore (NCT06341803), from March 2024 to March 2025. Ethical approval was obtained (DSRB 2023/00397). Inclusion criteria were: (1) age ≥ 21; (2) DSM-5 diagnosis of MDD with current major depressive episode; (3) baseline MADRS ≥ 20; and (4) at least one failed antidepressant trial. Exclusion criteria included psychosis, recent substance abuse, acute suicidality requiring inpatient care, significant neurological disorders (e.g. epilepsy), pregnancy, or contraindications to magnetic resonance imaging (MRI) or transcranial magnetic stimulation (TMS). Participants maintained stable medications during the treatment week.

Given the pilot design, the institutional review board (IRB) application included an approximate G*Power calculation indicating a minimum of 20 participants with paired pre-post observations, based on a large within-participant effect (Cohen’s *d* = 0.8; paired-sample t-test; α = 0.05). Under these assumptions, this sample size provides approximately 92.4% power to detect the assumed effect using a paired two-tailed t-test. Twenty-one participants were enrolled; one withdrew during treatment, leaving 20 participants with immediate post-treatment data. Because the calculation was based on continuous pre-post change rather than the binary primary response endpoint, it did not constitute a formal power calculation for the primary endpoint. The study was not powered for the subgroup, neuroimaging, or health-economic analyses.

### TMS treatment protocol

Stimulation was delivered with a MagPro X100 stimulator (MagVenture, Farum, Denmark). To ensure precise and reproducible coil placement, we utilized the Axilum Robotics TMS-Cobot (Axilum Robotics, Strasbourg, France) with the Localite neuronavigation system (Localite GmbH, Sankt Augustin, Germany). Real-time neuronavigation was maintained to keep the coil within 2 mm of the personalized target.

We employed the Stanford Accelerated Intelligent Neuromodulation Therapy (SNT) stimulation protocol using our TAO-TMS targets. Each session delivered 1800 pulses consisting of bursts of 3 pulses at 50 Hz, repeated at 5 Hz, with a train duration of 2 seconds and an inter-train interval of 8 seconds. The coil (MagVenture Cool-B65) was positioned tangential to the scalp with the handle oriented at 45 degrees to the mid-sagittal line. Participants received 10 sessions per day over 5 consecutive days (50 sessions total). Sessions were separated by a 50-minute inter-session interval.

Stimulation was delivered at 90% resting motor threshold (RMT), linearly adjusted for the cortical depth of the target relative to the motor hotspot (Stokes et al., 2005; Cole et al., 2020). To mitigate scalp discomfort, intensity was titrated from 70% to 90% of the adjusted RMT over the initial three sessions. Participants were asked at the end of each treatment session whether they had experienced any side effects or discomfort. Although a standardized adverse-event instrument was not administered, reported events were documented by the clinical team.

Treatment-delivery accuracy was quantified in each participant’s native T1 volumetric space. The Localite neuronavigation system recorded the coil position for each delivered pulse relative to the participant’s native MRI. These records were used to derive the actual treatment location for each pulse, and the Euclidean distance between this location and the intended TAO-TMS target was calculated in the individual’s native T1 space. Distances were averaged across all delivered pulses within a treatment session to obtain the mean spatial deviation (“COBOT accuracy”) for each participant.

### Clinical assessments and outcomes

Participants were assessed at baseline, post-treatment, 1-month and 3-month follow-ups using the MADRS score (Montgomery and Åsberg, 1979), Quick Inventory of Depressive Symptomatology (QIDS-SR16; Rush et al., 2003), Generalized Anxiety Disorder-7 (GAD-7; Spitzer et al., 2006), Montreal Cognitive Assessment (MoCA; Nasreddine et al., 2005), Sheehan Disability Scale (SDS; Sheehan et al., 1996), EuroQol-5D (EQ-5D; EuroQol Group, 1990), and the Quality of Life Enjoyment and Satisfaction Questionnaire (Q-LES-Q-SF; Endicott et al., 1993). QIDS-SR16 and GAD-7 scores were additionally assessed after each treatment day to capture short-term symptom changes. Clinician-rated assessments were administered by trained raters, and self-report measures were completed by participants.

The IRB-approved study application specified the primary endpoint as response immediately after treatment, defined as a ≥50% reduction from baseline in MADRS total score. QIDS-SR16, MoCA, EQ-5D, and Q-LES-Q-SF were prespecified secondary outcome measures. The remaining clinical outcomes were exploratory: MADRS response at the 1- and 3-month follow-ups and at any assessment within 4 weeks after treatment; remission (MADRS <11) immediately after treatment, at the 1- and 3-month follow-ups, and at any assessment within 4 weeks; and changes over time in GAD-7 and SDS scores. Analytic methods are described in *Statistical analysis*.

### MRI acquisition & preprocessing

All MRI scans were acquired at the Centre for Translational Magnetic Resonance Research (TMR), National University of Singapore, on a 3T Siemens Prisma scanner. For each participant, a T1-weighted structural scan and two 10-minute resting-state fMRI (rs-fMRI) runs were collected prior to treatment. Acquisition details are provided in Supplementary Methods. The scalp-based BeamF3 (Beam et al., 2009) stimulation location was also identified and marked on the scalp using a vitamin E capsule. Participants completed a second MRI session on average 5.0 ± 1.9 days after treatment, consisting of one T1-weighted structural scan and two resting-state fMRI runs. Data was preprocessed using the Computational Brain Imaging Group (CBIG) fMRI preprocessing pipeline (Li et al., 2019; Ooi et al., 2025; Kong et al., 2026). More details can be found in Supplementary Methods.

### Personalized TAO-TMS target

A personalized TAO-TMS target in the left dorsolateral prefrontal cortex (DLPFC) was generated for each participant using baseline rs-fMRI. To preserve individual anatomical fidelity, localization was performed in the participant’s native volumetric space. We employed the Multi-Session Hierarchical Bayesian Model (MS-HBM) to estimate reliable individual-specific cortical networks (Kong et al., 2019), which were then mapped into the participant’s native volumetric space. A tree-based algorithm was then used to estimate a personalized target within attention networks in left DLPFC that exhibited the strongest negative FC (based on Pearson’s correlation) with the subgenual anterior cingulate cortex (sACC), while remaining close to the scalp to minimize TMS intensity requirements (Kong et al., 2026). The attention networks were chosen because dorsal attention and salience/ventral attention networks are known to be negatively correlated with the default network (Fox et al., 2006) and thus the sACC time course (Siddiqi et al., 2023). We also note that the salience network has been shown to be enlarged in depression patients (Lynch et al., 2024).

To quantify the spatial variability of personalized TAO-TMS targets across participants, each participant’s target was transformed into every other participant’s native T1 space using Advanced Normalization Tools (ANTs; Avants et al., 2011). For each pair of participants, we computed the Euclidean distance between participant A’s transformed target and participant B’s target in participant B’s native space, and vice versa in participant A’s native space. These two distances were averaged to obtain a symmetric pairwise estimate. The mean pairwise distance was then calculated across all participant pairs. To compare personalized TAO-TMS targets with conventional BeamF3 targets, both targets were identified in each participant’s native T1 space, and their Euclidean distance was calculated directly within that space.

### Historical BeamF3 reference

To provide descriptive local context, we analyzed data from a previously published retrospective sample of patients who received non-accelerated BeamF3 TMS at the same hospital (N = 23; 49 ± 29 treatment sessions; mean number of comorbid psychiatric diagnoses = 1.2; Ye et al., 2024). Clinical improvement was quantified as the percentage reduction in MADRS score from baseline to immediately after treatment. As an exploratory analysis, to reduce baseline differences between the historical BeamF3 and TAO-TMS samples, propensity scores were estimated using logistic regression based on age, sex, baseline MADRS score, and number of comorbid psychiatric diagnoses. Using 1:1 nearest-neighbor matching with replacement, each TAO-TMS participant was matched to the historical BeamF3 participant with the closest propensity score. The resulting matched historical sample is referred to as “BeamF3 Ye2024 (PSM).” Given that participants were not randomized to BeamF3 or TAO-TMS targets, this analysis was intended to provide descriptive context and does not establish comparative efficacy.

### TMS-induced electric field

To characterize differences in the modeled electric field (E-field) distributions produced by TAO-TMS and BeamF3 (Beam et al., 2009) targets, individualized head models were generated from the anatomical MRI scans using SimNIBS 4.1.0 (Thielscher et al., 2015). For each participant, we simulated the E-field distribution for both the administered personalized target and the BeamF3 target (derived from the MRI-visible fiducial marker) using the MagVenture Cool-B65 coil model with a coil angle of 45 degrees. For each simulation, the “E-field hotspot” was defined as the set of cortical vertices representing the top 1% of E-field magnitude (Lynch et al., 2022; Ren et al., 2025).

Two descriptive metrics were calculated. First, network engagement was defined as the percentage of the E-field hotspot surface area falling within each of the 17 individualized cortical networks. Second, baseline hotspot sACC FC was calculated by correlating the sACC time course with an E-field-weighted average of DLPFC vertex time courses (Elbau et al., 2023). More negative values indicate stronger sACC anticorrelation and may indicate better target quality (Weigand et al., 2018; Elbau et al., 2023). However, because TAO-TMS was designed around both characteristics whereas BeamF3 was not, their comparison characterized modeled network engagement rather than establishing superior target quality or clinical efficacy.

### Treatment-related FC changes

Treatment-related functional connectivity changes were examined in participants who completed a post-treatment scan with acceptable head motion. The difference in hotspot sACC FC (post-treatment minus baseline) was correlated with the percentage improvement in MADRS scores. To examine whole-brain FC changes, for each participant, two 419 x 419 FC matrices were computed – one at baseline and one post-treatment – with 400 cortical (Schaefer et al., 2018) and 19 subcortical (Fischl, 2012) regions. Differences between the two matrices were then computed within each participant using xDF (Afyouni et al., 2019).

### Exploratory analyses of coil orientation and target constraints on clinical improvement

To explore whether coil orientation and targeting constraints were associated with clinical improvement, we conducted two sets of exploratory analyses. First, the coil orientation in our trial was fixed at 45 degrees, following the SNT stimulation protocol. For each participant, we modeled alternative coil orientations to minimize hotspot sACC FC (i.e., maximize anticorrelations). The difference between the minimum hotspot sACC FC and the actual hotspot sACC FC (based on the 45-degree coil angle) was then correlated with percentage MADRS improvement across participants. A positive correlation would suggest that optimizing coil orientation to further minimize hotspot sACC FC might potentially improve clinical outcomes. We repeated the same analysis by replacing hotspot sACC FC with attentional-network engagement. Here, a negative correlation would suggest that optimizing coil orientation to further maximize attention-network engagement might potentially improve clinical outcomes.

Second, recall that TAO-TMS restricted the target to be near the scalp and to be within the attention networks. To explore the impact of these targeting constraints, personalized TMS targets were regenerated without the near-scalp constraint, and the Euclidean distance between these unconstrained targets and the original targets was correlated with percentage MADRS improvement. A negative correlation would suggest that the near-scalp constraint was potentially worsening clinical outcomes. The same analysis was repeated by removing both the near-scalp and attentional network constraints.

### Exploratory health economic analysis

We conducted an exploratory cost-effectiveness analysis comparing TAO-TMS with ECT using a standard hybrid decision tree-Markov model over a one-year time horizon (Figure S1; Sonnenberg and Beck, 1993). Analyses were performed from healthcare system and societal perspectives.

The model comprised two stages with the same structure for TAO-TMS and ECT. First, the decision tree classified patients immediately after treatment into one of three acute-outcome categories: responders (≥50% reduction in MADRS), partial responders (≥25% but <50% reduction in MADRS), or nonresponders (<25% reduction in MADRS). Second, at 1 month after treatment, the same patients entered the Markov model and were assigned to one of three health states based on their MADRS score at that assessment: remission (MADRS <11), partial relapse (MADRS 11-20), or relapse (MADRS >20). Thus, the acute-outcome categories described the initial percentage improvement, whereas the Markov health states described symptom severity at 1 month. During each subsequent monthly cycle through month 12, patients could remain in their current health state or transition to another state.

Treatment-specific probabilities were assigned to this common model structure. For TAO-TMS, data from the present trial were used to determine both the probabilities of the acute-outcome categories and the probabilities of moving from each acute-outcome category immediately after treatment to each health state at 1 month after treatment (when the Markov model began). Subsequent transition probabilities among the health states were estimated from changes observed between the 1-month and 3-month assessments. Because these assessments were separated by two months, the observed transition probabilities were converted into monthly probabilities assuming constant transition hazards. These monthly probabilities were then applied through month 12.

For ECT, the corresponding acute-outcome and follow-up probabilities were obtained from published economic evaluations of nonpsychotic TRD populations (Zhao et al., 2018; Fitzgibbon et al., 2020). Because TAO-TMS outcomes were unavailable beyond 3 months and the published ECT inputs did not include recovery from relapse, relapse was treated as an absorbing state in both arms: once patients entered relapse, they remained in that state for the remainder of the modeled period.

Health-state utility values were derived from EQ-5D data collected in the present trial. Quality-adjusted life-years (QALYs) were calculated by weighting the time spent in each health state by its corresponding utility value (Drummond et al., 2015). Costs included TMS, ECT, imaging, medication, adverse events, and productivity loss. Cost inputs were drawn from published Singapore Ministry of Health rates, Singapore Agency for Care Effectiveness resource sheets, institutional cost data, and data from the present trial. All model parameters and their sources are reported in Table S6.

Incremental cost-effectiveness ratios (ICERs) were calculated (Weinstein et al., 1996), and uncertainty was assessed using one-way sensitivity analyses and probabilistic sensitivity analyses with 10,000 Monte Carlo simulations (Briggs, 1999; Briggs et al., 2006). From these simulations, cost-effectiveness acceptability curves (CEACs) were generated to show the probability that TAO-TMS is more cost-effective than ECT across a range of willingness-to-pay thresholds. Scenario analyses examined shorter time horizons (3-month and 6-month) and two health states (instead of three). Additional details are provided in the Supplementary Methods *Exploratory health economic analysis*. All costs were analyzed in 2025 Singapore Dollars (SGD) and also presented in US Dollars (USD) using a fixed exchange rate of 1 USD = 1.29 SGD.

### Statistical analysis

#### Clinical outcomes

All clinical outcome analyses were conducted using SPSS version 32 (IBM, Armonk, NY). Analyses included participants with available data at each assessment, and missing data were not imputed. The primary endpoint – MADRS response immediately after treatment – and the exploratory response and remission outcomes were summarized as proportions with exact Clopper-Pearson 95% confidence intervals.

Longitudinal changes in continuous outcomes were examined using separate linear mixed-effects models, with time specified as a categorical fixed effect and participant as a random intercept. No additional covariates were included. Denominator degrees of freedom were estimated using the Satterthwaite approximation. Models were estimated using all available observations. All pairwise contrasts among assessment timepoints were derived from estimated marginal means and Bonferroni-corrected within each measure. Daily QIDS-SR16 and GAD-7 scores were analyzed using the same model with day (baseline and treatment days 1-5) as the fixed effect, with all pairwise contrasts among days Bonferroni-corrected within each measure. Model residuals were screened for normality using the Shapiro-Wilk test, and no departures warranting rank-based alternatives were identified. All tests were two-sided with α = 0.05.

#### Neuroimaging and exploratory analyses

Analyses were conducted using in-house MATLAB (2018, MathWorks, Natick, MA) and Python (3.9.12) scripts. E-field characteristics (network specificity and hotspot sACC FC) of TAO-TMS and BeamF3 targets were compared based on the 20 participants who completed all 50 TMS sessions with paired two-tailed t-tests. Treatment-related functional connectivity changes were examined in 18 participants who completed a post-treatment scan with acceptable head motion. Within-participant whole-brain FC change between baseline and post-treatment was quantified using xDF (Afyouni et al., 2019), which accounts for temporal autocorrelation in fMRI time series and yields an edge-wise z-statistic for FC change (post-treatment minus baseline) in each individual. Multiple comparisons were corrected within each individual with a false discovery rate of q < 0.05. The association between hotspot sACC FC change and MADRS improvement was assessed using Pearson’s correlation. Exploratory analyses of coil orientation and target constraints used Pearson’s correlations and paired two-tailed t-tests. These analyses were not prespecified and are interpreted as hypothesis-generating. For each set of analyses, multiple comparisons were corrected with a false discovery rate of q < 0.05.

#### Health economic evaluation

Cost-effectiveness was assessed using a hybrid decision tree-Markov model with one-way and probabilistic sensitivity analyses, as described in *Exploratory health economic analysis*. Analyses were performed in RStudio Desktop Pro v2024.04.1 (Posit Software, PBC, Boston, MA), with modeling conducted in TreeAge Pro 2023 R2 (TreeAge Software, LLC, Williamstown, MA).

### Code availability

The fMRI preprocessing pipeline is available at https://github.com/ThomasYeoLab/CBIG/tree/master/stable_projects/preprocessing/CBIG_fMRI_Preproc2016. The individual-specific MS-HBM parcellation code is available at https://github.com/ThomasYeoLab/CBIG/tree/master/stable_projects/brain_parcellation/Kong20 19_MSHBM. The TAO-TMS targeting code is available at https://github.com/ThomasYeoLab/Kong2026_TMSTree.

## Results

### Study population and safety

Figure 1 shows the CONSORT flow diagram. Of 27 participants screened, 21 participants were recruited (mean age 35.1 ± 12.3 years; 57% female; 81% Chinese, 19% South Asian). The sample exhibited substantial clinical complexity (Table 1), with 86% meeting criteria for at least one additional psychiatric diagnosis; the average number of comorbid psychiatric diagnoses was 1.67. The enrolled participants had 4.7 ± 2.3 failed antidepressant trials, and 19% had previously received electroconvulsive therapy (ECT). Mean baseline MADRS was 28.3 ± 5.1. Detailed demographic and clinical characteristics are summarized in Table S1.

**Figure 1.**
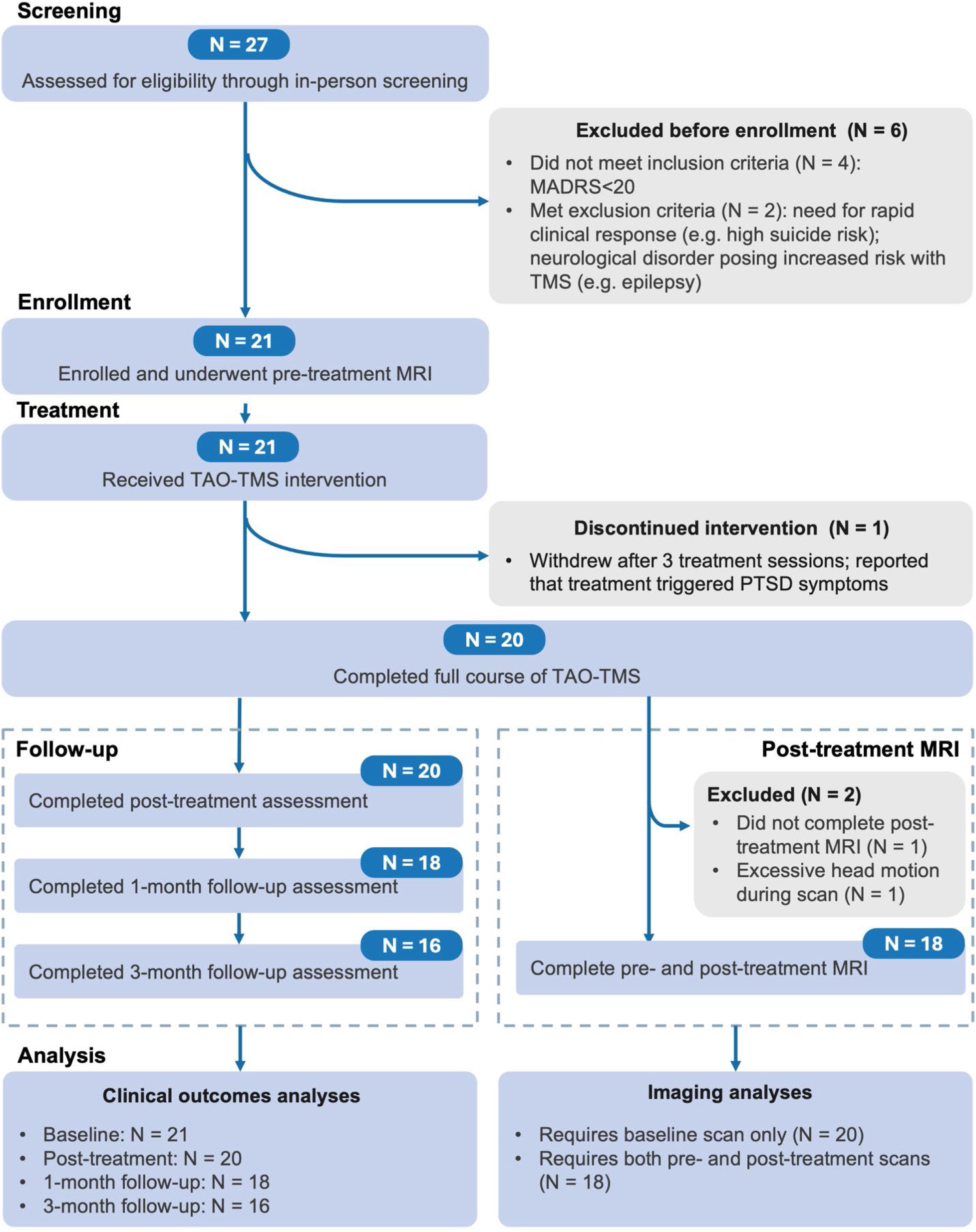
CONSORT flow diagram. TMS, transcranial magnetic stimulation; MADRS, Montgomery-Asberg Depression Rating Scale; PTSD, post-traumatic stress disorder; MRI, magnetic resonance imaging.

**Table 1.** Baseline demographic and clinical characteristics of all intent-to-treat participants.

| Baseline characteristic | Participants (N=21) |
|---|---|
| Age, years, mean (SD) | 35.14 (12.25) |
| Gender, No. (%) |  |
| Male | 9 (43) |
| Female | 12 (57) |
| Ethnicity, No. (%) |  |
| Chinese | 17 (81) |
| South Asian | 4 (19) |
| Comorbidities, No. (%) |  |
| Generalized anxiety disorder | 12 (57) |
| Post-traumatic stress disorder | 6 (29) |
| Personality disorder | 6 (29) |
| Feeding and eating disorder | 5 (24) |
| Autism spectrum disorder | 3 (14) |
| Bipolar II disorder | 1 (5) |
| Attention-deficit/hyperactivity disorder | 1 (5) |
| Obsessive-compulsive disorder | 1 (5) |
| Gender dysphoria with gender-affirming surgery | 1 (5) |
| Breast and kidney cancer | 1 (5) |
| Hepatitis B | 1 (5) |
| Number of failed antidepressant trials, mean (SD) | 4.67 (2.27) |
| Past ECT treatment, No. (%) | 4 (19) |
| Past TMS treatment, No. (%) | 8 (38) |
| Clinical assessments, mean (SD) |  |
| MADRS | 28.33 (5.14) |
| QIDS-SR16 | 20.67 (6.45) |
| GAD-7 | 10.86 (5.26) |
| Q-LES-Q-SF | 39.00 (9.41) |
| EQ-5D-3L utility score | 0.39 (0.33) |
| SDS total score | 17.86 (6.46) |
| MoCA | 28.52 (1.47) |

ECT, electroconvulsive therapy; TMS, transcranial magnetic stimulation; MADRS, Montgomery-Asberg Depression Rating Scale; QIDS-SR16, The Quick Inventory of Depressive Symptomatology (16-Item) (Self-Report); GAD-7, 7-item Generalized Anxiety Disorder scale; Q-LES-Q-SF, Quality of Life Enjoyment and Satisfaction Questionnaire-Short form; EQ-5D-3L, EuroQol 5-dimensional questionnaire with 3 levels; SDS, Sheehan Disability Scale; MoCA, the Montreal Cognitive Assessment.

Of the 21 enrolled participants, 20 completed the full 50-session treatment protocol (95.2%). Ten participants (47.6%) reported at least one adverse event, whereas 11 (52.4%) reported none (Table S2). The most commonly reported events were treatment-site discomfort or headache during stimulation (5/21; 23.8%), insomnia (4/21; 19.0%), and anxiety (3/21; 14.3%). One participant (4.8%) withdrew after three treatment sessions, reporting that treatment had triggered PTSD symptoms. No serious adverse events were reported.

The final treated cohort had a mean age of 34.9 ± 12.5 years with mean baseline MADRS of 28.4 ± 5.3 and 4.7 ± 2.3 failed antidepressant trials. The average number of comorbid psychiatric diagnoses was 1.6, and only three participants had no comorbid psychiatric diagnosis. Overall, the 95.2% treatment-completion rate supports the tolerability and deliverability of the robotic TAO-TMS protocol in this population.

### Personalized TAO-TMS targets

TAO-TMS was used to derive a personalized target in each participant’s native T1 volumetric space using baseline rs-fMRI. Figure 2A illustrates the spatial distribution of personalized targets overlaid on group-level cortical networks (Yeo et al., 2011; Kong et al., 2019). Participants exhibited highly distinct network topography (Figure 2B top panel). All TAO-TMS target locations were close to the scalp (Figure 2B bottom panel) with an average distance of 14.1 mm from the scalp as measured in native volumetric space. The mean pairwise distance between the personalized targets across the 20 participants was 9.6 mm (Figure 2C). For context, the mean distance between the personalized TAO-TMS targets and the traditional scalp-based BeamF3 targets was 16.1 mm (Figure 2D), with a maximum deviation of 32.1 mm.

**Figure 2.**
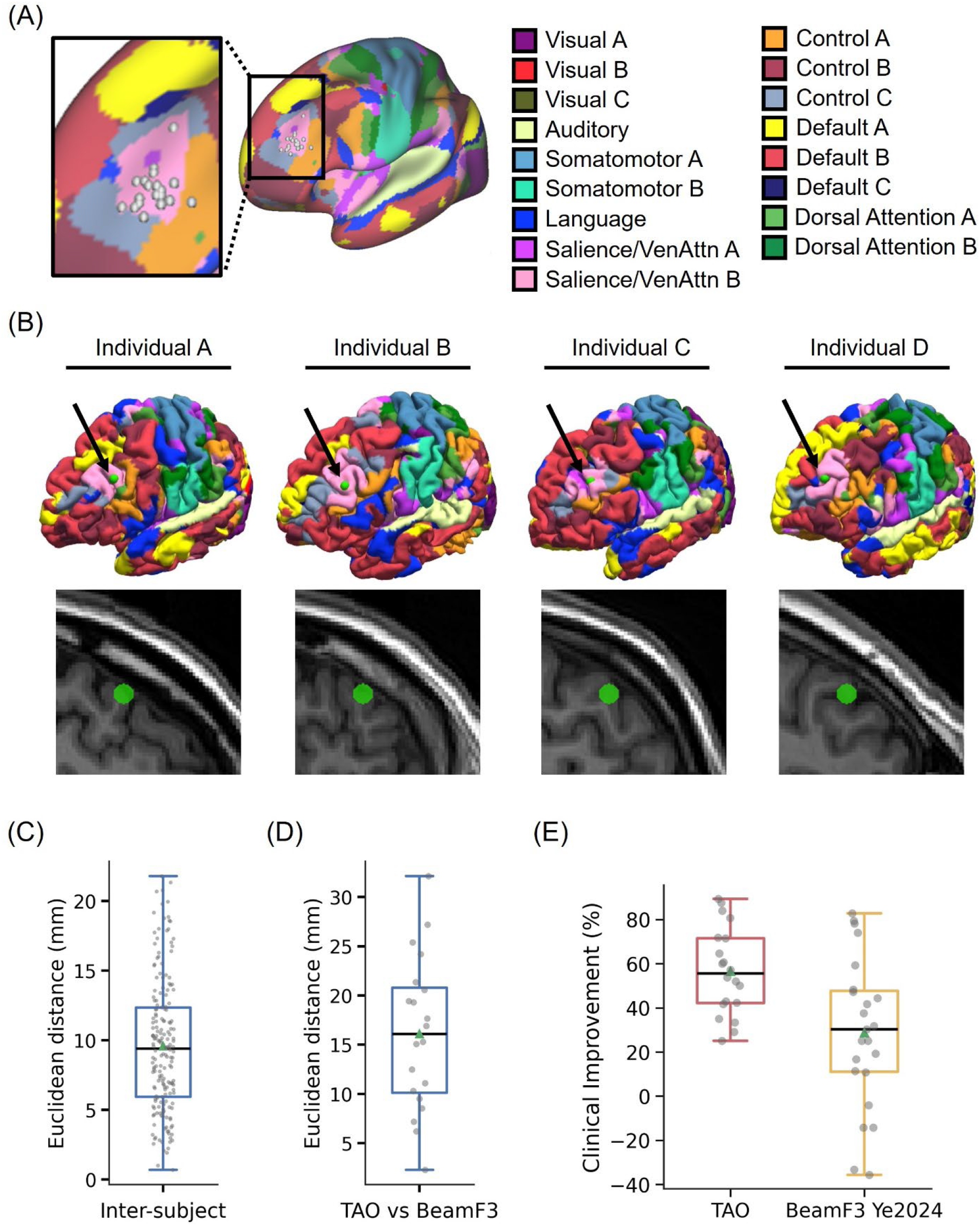
Comparison of TAO-TMS and BeamF3 targets. (A) Personalized depression targets of 20 participants overlaid on a group-level map of 17 resting-state networks (Yeo et al., 2011; Kong et al., 2019). Target locations were estimated in each participant’s native T1 volumetric space using baseline rs-fMRI and then projected to fsaverage6 surface space for visualization. (B) TAO-TMS targets are situated within personalized attention networks and are located close to the scalp. Four representative participants are shown here, with targets (green circles) displayed on native pial surfaces (top) and native T1 images (bottom). Black arrows point to the green circles on the native pial surfaces. A target located deep within a sulcus would not be visible on the native pial surface, so the visible targets (green circles) indicate that TAO-TMS was able to successfully localize near-scalp targets. (C) Boxplot of the pairwise distances between the personalized targets across the 20 participants. (D) Boxplot of the distances between the personalized targets and the scalp-based BeamF3 targets across 20 participants. (E) Clinical improvement with TAO-TMS (N = 20) and with non-accelerated BeamF3 TMS in a retrospective sample from the same hospital (N = 23; # sessions = 49 ± 29; Ye et al., 2024). Boxplots show percentage reduction in MADRS scores immediately after treatment. Positive values indicate greater improvement.

### Clinical outcomes of TAO-TMS

To provide descriptive local context, Figure 2E presents the mean MADRS reduction in the TAO-TMS sample (57% ± 19%; N = 20) alongside that observed in a retrospective sample receiving non-accelerated BeamF3 TMS at the same hospital (29% ± 34%; N = 23; Figure 2E; Ye et al., 2024). The historical sample received 49 ± 29 TMS sessions and had a mean of 1.2 comorbid psychiatric diagnoses. An exploratory propensity-matched analysis of the historical sample achieved a MADRS reduction of 19% ± 45% (Figure S2). Because the samples differed in study design, treatment schedule, and clinical characteristics, the historical findings are presented only as descriptive context and cannot establish comparative efficacy.

Within the TAO-TMS sample, the primary endpoint – the MADRS response rate immediately after treatment – was 65% (95% CI, 40.8%-84.6%; Table S3). In exploratory analyses, response rates were 50% (95% CI, 26.0%-74.0%) at 1 month and 31.3% (95% CI, 11.0%-58.7%) at 3 months. The corresponding remission rates were 30% (95% CI, 11.9%-54.3%), 27.8% (95% CI, 9.7%-53.5%), and 6.3% (95% CI, 0.2%-30.2%), respectively (Table S3). At any assessment within 4 weeks after treatment, 70% of participants achieved response (95% CI, 45.7%-88.1%) and 45% achieved remission (95% CI, 23.1%-68.5%).

In exploratory longitudinal analyses, depressive symptoms assessed with the MADRS changed significantly over time (*F*(3, 52.33) = 23.69, *p* < 0.001) and were lower than baseline immediately after treatment (mean difference = 16.24, 95% CI 10.89 to 21.58, *p* < 0.001), at 1 month (mean difference = 10.11, 95% CI 4.59 to 15.63, *p* < 0.001), and at 3 months (mean difference = 7.98, 95% CI 2.25 to 13.70, *p* = 0.002; Figure 3A; Table S4). Self-reported symptoms of depression (QIDS-SR16) and anxiety (GAD-7) also changed significantly over time (QIDS-SR16: *F*(3, 58.36) = 11.46, *p* < 0.001; GAD-7: *F*(3, 51.94) = 7.36, *p* < 0.001). QIDS-SR16 scores were lower immediately after treatment and at 1 month, but not at 3 months, whereas GAD-7 scores were lower only immediately after treatment. Both measures also decreased across the five treatment days (QIDS-SR16: *F*(5, 95.83) = 18.55, *p* < 0.001; GAD-7: *F*(5, 95.39) = 17.07, *p* < 0.001; Figure S3; Tables S4 and S5).

**Figure 3.**
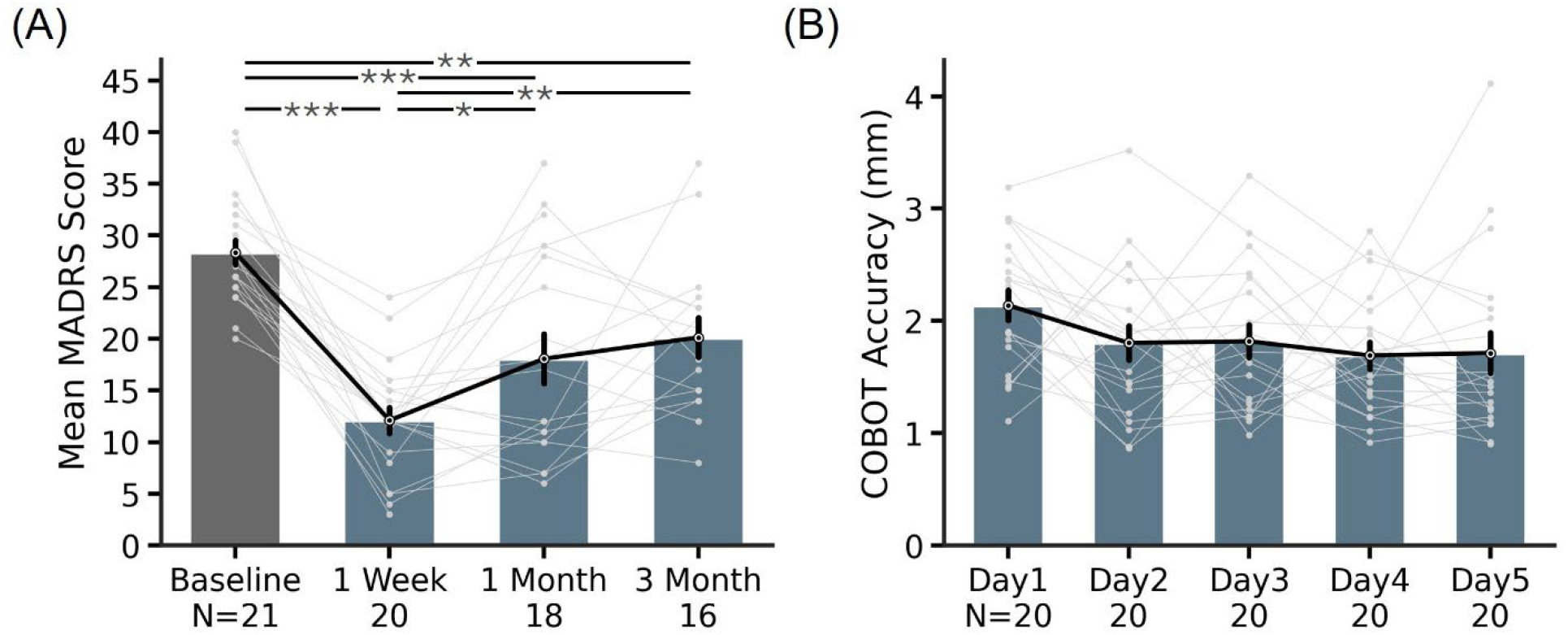
Clinical outcomes and treatment delivery accuracy. (A) Mean MADRS scores at baseline, post-acute treatment (1 week), 1-month, and 3-month follow-up. * indicates statistical significance after Bonferroni correction for multiple comparisons. *p<0.05. **p<0.01. ***p<0.001. (B) COBOT accuracy for personalized TAO-TMS target delivery across the five treatment days, expressed as the mean Euclidean distance between the actual stimulation site and the planned stimulation site. Error bars represent standard error.

Functional impairment (SDS) and enjoyment and satisfaction with life (Q-LES-Q-SF) changed significantly over time (SDS: *F*(3, 52.07) = 6.82, *p* < 0.001; Q-LES-Q-SF: *F*(3, 51.83) = 5.96, *p* = 0.001), with improvements immediately after treatment and at 1 month, but not at 3 months. General health-related quality of life (EQ-5D utility) increased from 0.39 at baseline to 0.60 immediately after treatment, although the overall effect of time was not significant (*F*(3, 53.04) = 2.56, *p* = 0.065). Global cognitive performance (MoCA) showed an overall effect of time (*F*(3, 51.63) = 3.45, *p* = 0.023), although no post-treatment assessment differed significantly from baseline. Full descriptive statistics and pairwise contrasts are reported in Tables S3-S5. Reported pairwise *p* values were Bonferroni-corrected within each measure.

Throughout the treatment course, stimulation was delivered reliably to the personalized targets, with a mean deviation of 1.8 ± 0.7 mm from the intended target (Figure 3B).

### E-field characteristics of TAO-TMS and BeamF3 targets

TAO-TMS selects a personalized target within attention networks in left DLPFC that exhibits the strongest negative FC with sACC (Figure 4A). E-field modeling indicated that salience/ventral attention network B showed the greatest mean engagement within TAO-TMS hotspots, followed by control and default networks, while the other attention networks contributed relatively little (Figure 4B). Although the relative contributions varied substantially across individuals, engagement was generally concentrated within this limited set of networks (Figure 4C).

**Figure 4.**
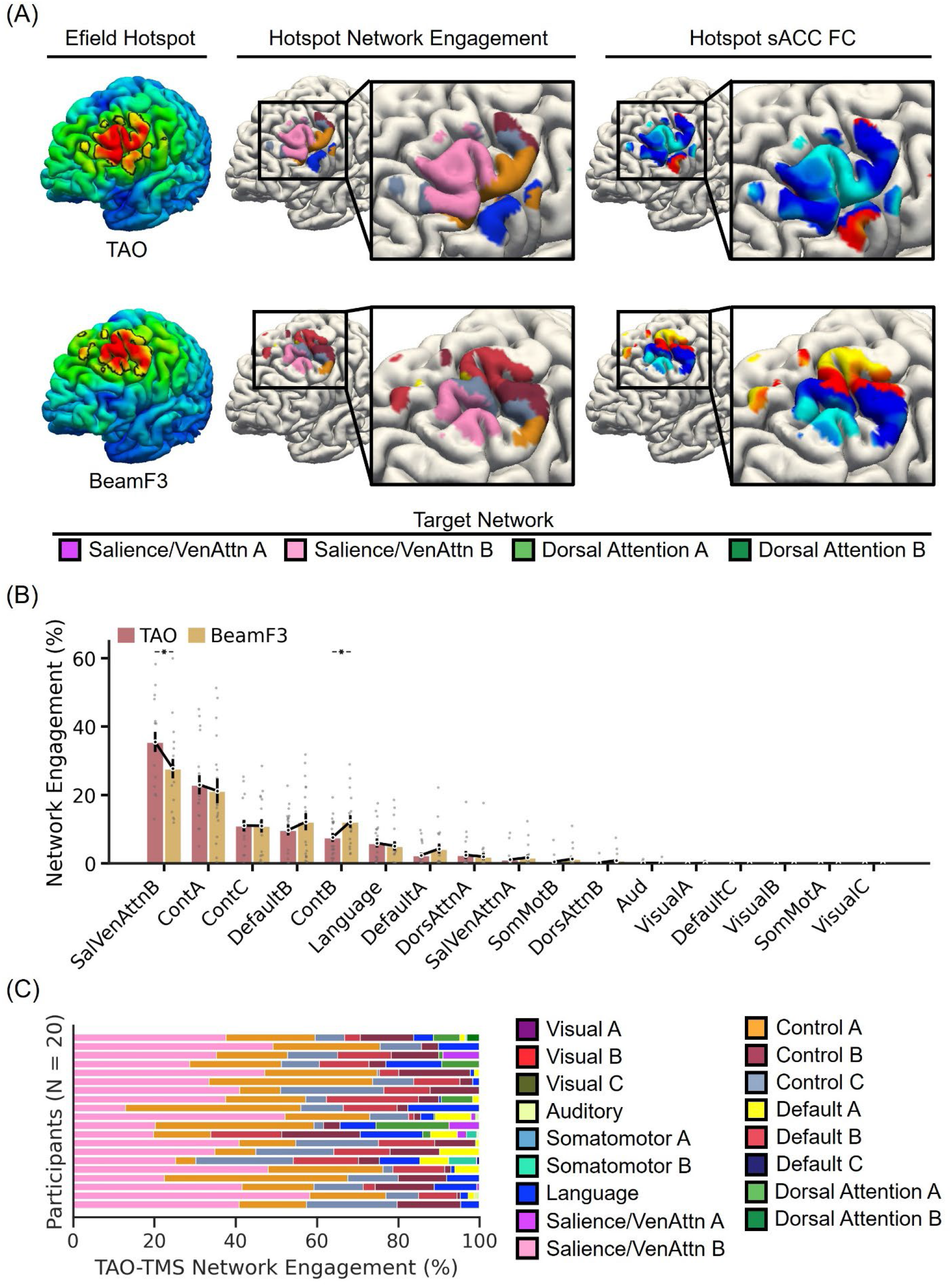
Electric field characteristics of TAO-TMS and BeamF3 targets. (A) E-field characteristics of the TAO-TMS target (top row) and the BeamF3 target (bottom row) in a representative participant. The left panel shows the E-field maps estimated using SimNIBS, with the “E-field hotspot” defined as the top 1 percent of cortical vertices (black boundaries) following previous studies (Lynch et al., 2022; Ren et al., 2025). The middle panel shows the personalized MS-HBM networks within the E-field hotspot. The right panel shows functional connectivity between the E-field hotspot and the sACC. Target networks are the salience/ventral attention networks A and B and dorsal attention networks A and B. (B) Network engagement of E-field hotspots for TAO-TMS and BeamF3 targets across large-scale cortical networks (Yeo et al., 2011; Kong et al., 2019) in 20 participants at baseline. * indicates statistical significance after FDR correction (q < 0.05). (C) Distribution of TAO-TMS E-field hotspot engagement across individual-specific networks for the 20 individual participants.

Despite BeamF3 not being designed to target attention networks, the overall ordering of network engagement was broadly similar between TAO-TMS and BeamF3 (Figure 4B). As expected from the targeting design, TAO-TMS hotspots showed 21.4% stronger engagement of attention networks (*p* = 0.012; Figure S4A) and 87.6% stronger (more negative) sACC FC (*p* = 0.012; Figure S4B) than BeamF3 targets at baseline. These results confirm that TAO-TMS achieved its intended targeting characteristics but do not establish clinical superiority.

### Treatment-related functional connectivity changes

Of the 20 participants who completed treatment, one missed the post-treatment scan and one was excluded due to excessive motion, leaving 18 participants for the following analyses. We investigated whether clinical improvement was related to FC changes from pre-treatment to post-treatment in 18 participants. Recall that the E-field hotspot (top row in Figure 4A) corresponded to the actual stimulation hotspot. There was no significant correlation between percentage MADRS reduction and change in hotspot sACC FC (*r* = -0.18, *p* = 0.476; Figure S4C).

At the whole-brain level, there were statistically significant changes in whole-brain FC in every participant (FDR q < 0.05), but the spatial distributions of these significant edges were highly variable across participants (Figure 5; Figures S5 to S7). Qualitatively, responders might exhibit greater FC changes than non-responders, but the between-group difference was not statistically significant. We note that among the non-responders, one participant displayed substantial FC changes post-treatment and subsequently showed clinical response at the 3-month follow-up (non-responder 4 in Figures S6 to S7).

**Figure 5.**
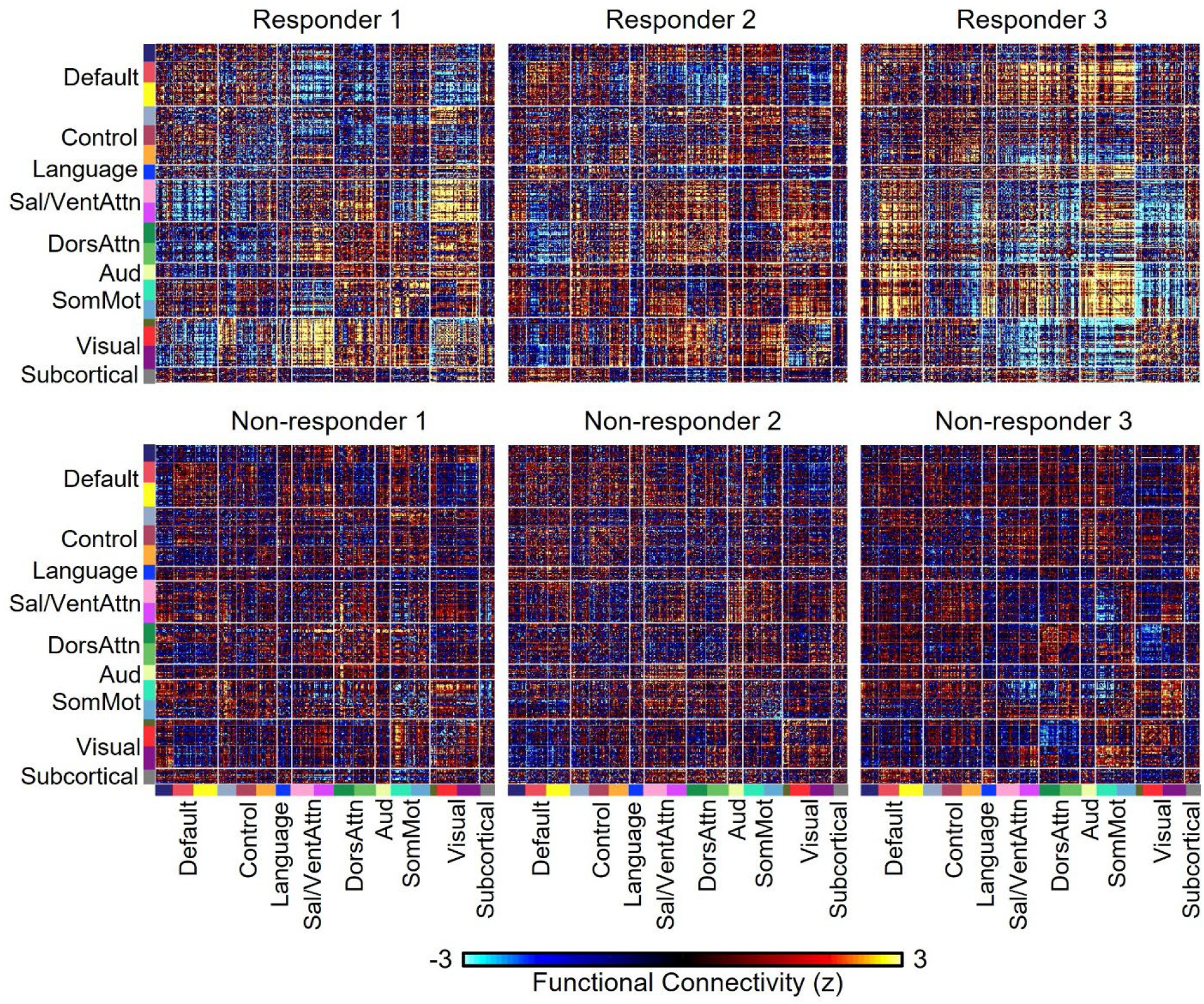
Functional connectivity (FC) changes from pre-treatment to post-treatment following TAO-TMS. Top panel shows three responders, while bottom panel shows three non-responders. Figure S5 shows thresholded FC changes after FDR correction (q < 0.05). FC changes for the remaining participants are shown in Figures S6 and S7.

### Impact of coil orientation & target constraints on observed clinical improvements

Following the SNT stimulation protocol, coil orientation was fixed at 45 degrees. In post hoc exploratory analyses, modeled changes in attention-network engagement and hotspot sACC FC under alternative coil orientations were not significantly associated with percentage MADRS improvement in this small sample (attention-network engagement: *r* = -0.35, *p* = 0.16; hotspot sACC FC: *r* = 0.14, *p* = 0.59; Figure S8).

TAO-TMS constrained targets to lie near the scalp and within the attention networks. In post hoc exploratory analyses, target displacement resulting from removal of these constraints was not significantly associated with percentage MADRS improvement in this small sample (removal of the near-scalp constraint alone: *r* = -0.18, *p* = 0.46, Figure 6A; removal of both the near-scalp and attention-network constraints: *r* = -0.08, *p* = 0.73, Figure 6B).

**Figure 6.**
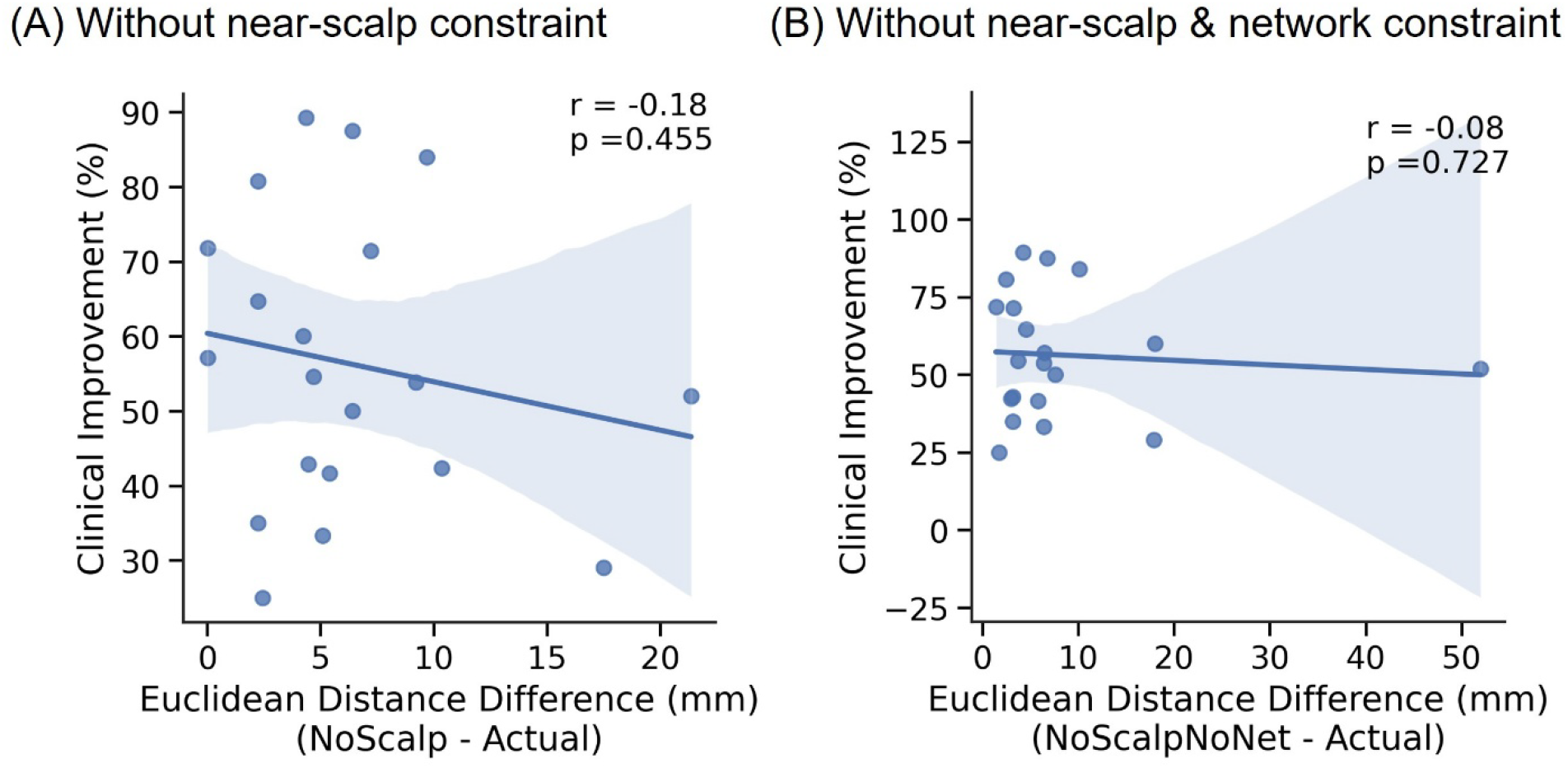
Post hoc exploratory analysis of TAO-TMS targeting constraints. (A) Correlation between percentage MADRS improvement and the distance from the actual TAO-TMS target to the personalized target generated without the near-scalp constraint. (B) Correlation between percentage MADRS improvement and the distance from the actual TAO-TMS targets to the personalized targets generated without the near-scalp and attentional-network constraints. Both correlations were directionally consistent with a possible clinical benefit of relaxing the targeting constraints, but neither reached statistical significance in this small sample.

### Exploratory health economic outcomes

In an exploratory cost-effectiveness analysis, the base-case model estimated lower costs and slightly higher quality-adjusted life-years (QALYs) for TAO-TMS than for ECT. From the healthcare system perspective, the estimated costs were S$45,172 (US$35,017) for TAO-TMS and S$55,767 (US$43,230) for ECT, corresponding to a difference of S$10,596 (US$8,214), while QALYs were similar: 0.67 (TAO-TMS) vs 0.65 (ECT). From the societal perspective, which included productivity losses, the model estimated a cost difference of S$19,213 (US$14,894) in favor of TAO-TMS (Table S7).

Cost-effectiveness acceptability curves (Figure 7) indicated a high model-estimated probability that TAO-TMS was cost-effective across the evaluated willingness-to-pay thresholds. In one-way sensitivity analyses, the conclusion from the healthcare system perspective was most sensitive to the modeled cost of ECT: TAO-TMS lost dominance when ECT treatment cost fell below S$695 (US$539). The societal perspective remained unchanged across parameter variations (Figure S9). Across shorter time horizons (3 and 6 months; Table S8) and the conservative two-state Markov model (Table S9; Figure S10), TAO-TMS remained less costly than ECT, while estimated QALYs were similar between treatments. Further details can be found in Supplementary Results.

**Figure 7.**
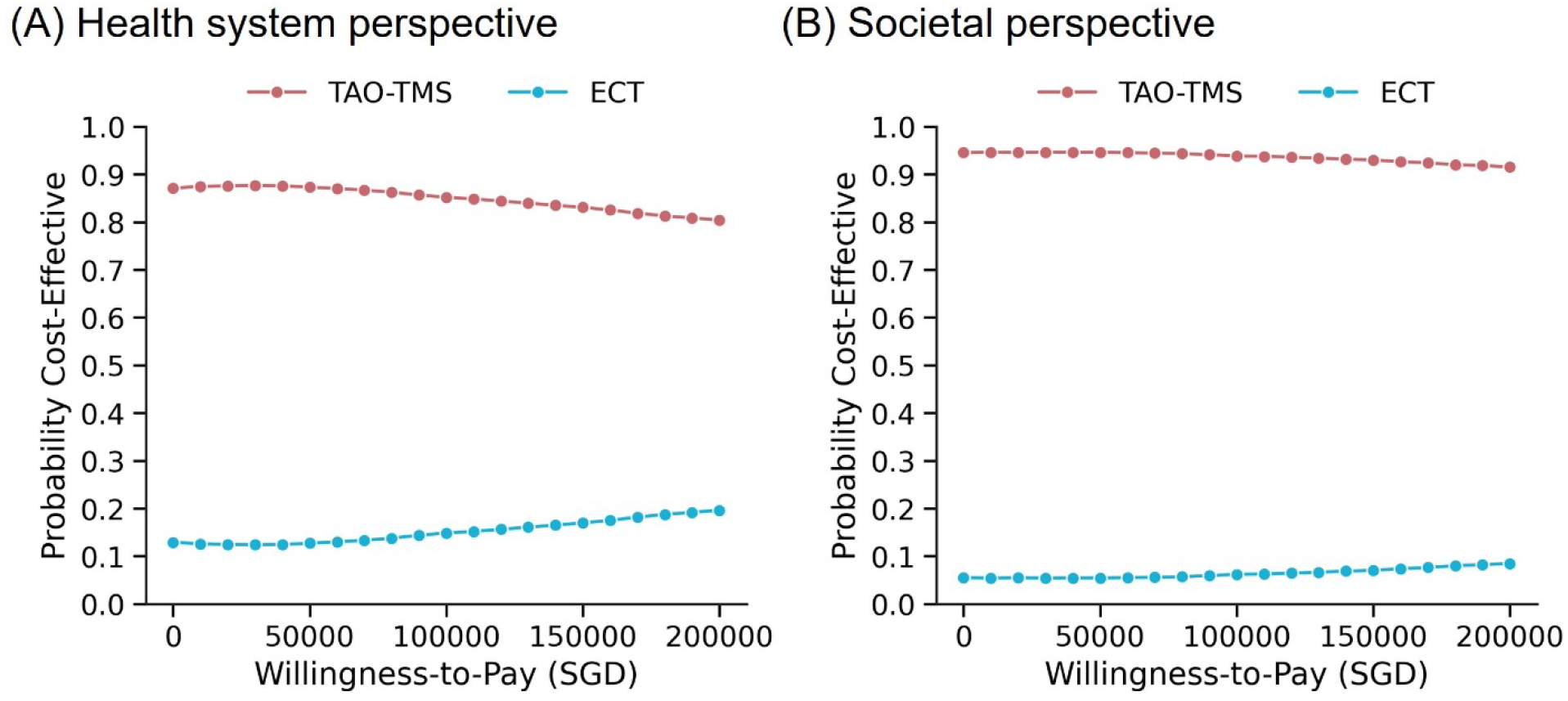
Exploratory cost-effectiveness acceptability curves for TAO-TMS versus ECT. The probability that TAO-TMS was cost-effective compared with ECT across a range of willingness-to-pay thresholds from (A) the healthcare system perspective and (B) the societal perspective, which includes productivity losses. The model estimated a high probability that TAO-TMS was cost-effective across the evaluated thresholds. Note that US$1 ≈ S$1.29.

## Discussion

This study supports the feasibility of personalized connectivity-guided accelerated iTBS in a naturalistic TRD population with complex psychiatric comorbidities often excluded from previous trials. TAO-TMS demonstrated a 65% response rate immediately after treatment, compared with the historical response rate of 22% with non-accelerated BeamF3 TMS from the same hospital (Ye et al., 2024). Post hoc E-field analysis confirmed that TAO-TMS achieved its intended targeting characteristics while revealing a broadly similar ordering of network engagement between TAO-TMS and BeamF3. Statistically significant post-treatment FC changes were found within each individual, but the study was not sufficiently powered to find any relationship between FC changes and treatment response. Finally, exploratory economic modeling estimated lower costs and similar QALYs for TAO-TMS relative to ECT.

### Effects of psychiatric comorbidities and target personalization on clinical outcomes

Psychiatric complexity may have contributed to our observed clinical outcomes. In accelerated iTBS trials using anatomical/scalp targets, naturalistic cohorts with substantial psychiatric comorbidity have reported response rates of 22%-28% (Ye et al., 2024; Goodman et al., 2025; Persson et al., 2025), compared with 52%-75% in studies applying more restrictive psychiatric eligibility criteria (Luehr et al., 2024; Zhao et al., 2024; Ramos et al., 2025). Although confounded by population and protocol differences, these informal cross-study comparisons suggest that psychiatric complexity may influence treatment response.

Stronger evidence comes from within-study comparisons of patients with and without complex comorbidities. Hearne et al. (2025) reported response rates of 74% in trial-eligible patients and 44% in patients with complex comorbidities following unaccelerated connectivity-guided iTBS. Consistently, when we excluded participants with autism, OCD and personality disorders from our TAO-TMS sample, the response rate in the remaining subgroup (N = 12) increased from 65% to 75% immediately after treatment and from 50% to 60% at 4 weeks among participants with available data, while remission was essentially unchanged. Although absolute response rates cannot be compared because the populations, protocols and assessment timing differed, the direction of the within-study contrasts was consistent, supporting the possibility that psychiatric comorbidity may contribute to lower treatment response.

Zhao et al. (2024) provides an informative comparison: the study’s iTBS arm delivered the same 90,000-pulse, five-day schedule using a predefined group-average left DLPFC target. Response rates were comparable with our study, but acute symptom reduction and remission were numerically higher in our study. However, Zhao et al. excluded participants with any secondary psychiatric disorder or prior ECT, whereas our participants averaged 1.7 psychiatric comorbidities and 19% had previously received ECT. The similar response rates could indicate no clinical advantage of individualized TAO-TMS targeting over Zhao’s group-average target or reflect differences in psychiatric complexity and prior treatment exposure. These explanations cannot be separated through cross-study comparison.

Targeting may also influence outcomes. Two recent randomized trials directly compared individualized connectivity-guided targeting with conventional scalp-based targeting for accelerated iTBS. Taylor et al. (2026a) found superior outcomes with individualized connectivity-guided targeting at one month, while Li et al. (2026) reported an advantage at week 6. However, Li et al. (2026) excluded participants with any secondary psychiatric disorder, while Taylor et al. (2026a) applied eligibility criteria similar to those of the SNT trials, which excluded several psychiatric comorbidities represented in our sample. These restrictive criteria leave uncertainty about the generalizability of their findings to more complex, naturalistic populations. Separately, Taylor et al. (2026b) found similar improvement in depressive symptoms following stimulation of two spatially distinct circuit-defined targets in patients with MDD and comorbid anxiety, whereas anxiety improved more with the anxiosomatic target. These findings suggest that multiple targets may be therapeutically meaningful for depression while differing in their effects on specific symptom domains.

Our prior work showed that TAO-TMS achieved greater reliability, closer scalp proximity and stronger sACC anticorrelation than several alternative individualized targeting approaches (Kong et al., 2026). Because the SNT targeting algorithm is proprietary, it was not included in that comparison. Reliability primarily reduces localization uncertainty, while scalp proximity may allow lower stimulation intensity and improve tolerability. sACC anticorrelation is an associative targeting marker whose causal and predictive validity remains under investigation. Although desirable, these target characteristics should not be interpreted as validated surrogates for clinical efficacy.

We cannot rule out that SNT targets are more clinically effective than TAO-TMS targets. Both the open-label and double-blind SNT trials reported substantially higher response and remission rates than the present study (Cole et al., 2020, 2022). However, both trials excluded complex psychiatric comorbidities, including autism, personality disorders and OCD. The active arm of Cole et al. (2022) averaged approximately 0.43 psychiatric comorbidities per participant, compared with 1.7 in our sample. More recently, Feyder et al. (2026) reported SNT outcomes with the same 5-day stimulation schedule in a larger, naturalistic multi-center sample. Although participants in Feyder et al. averaged approximately 1.0 psychiatric comorbidity versus 1.7 in our sample, acute MADRS reduction was 41.3% versus 57% in our study. These cross-study findings cannot establish relative effectiveness. Direct randomization within the same clinical population is required to compare targeting approaches.

### TMS-related functional connectivity

In our study, FC changes between the stimulation site and the sACC did not significantly correlate with symptom improvement. At the whole-brain level, statistically significant post-treatment FC changes were found within each participant. However, the spatial distribution of these changes varied markedly across individuals and was not associated with treatment response.

Gajawelli et al. (2024) found no significant active-versus-sham difference in FC change between individualized stimulation targets and the sACC. However, active SNT increased anticorrelation between the target and the default network relative to sham, and FC change between the target and a medial PFC region within the default network was associated with symptom improvement. More broadly, a recent review found mixed evidence regarding the relationship between TMS-related FC changes and symptom improvement (Briley et al., 2024). Some studies have reported significant associations (Philip et al., 2018; Eshel et al., 2020; Oathes et al., 2023), while other studies reported weak or non-significant associations (Ge et al., 2022; Stöhrmann et al., 2023; Jin et al., 2025).

The substantial heterogeneity observed in our study may partly reflect the complex psychiatric comorbidities in our sample. A larger sample size would be necessary to identify reproducible treatment-related FC changes and relate them to baseline psychiatric heterogeneity and clinical response.

### Health economics: policy and system-level implications

Under the base-case assumptions, exploratory health economic modeling estimated that TAO-TMS was less costly than ECT by approximately US$8,214 per patient from the healthcare system perspective and US$14,894 from the societal perspective, while QALYs were similar. TAO-TMS remained less costly, with similar QALYs, across shorter time horizons (3 and 6 months) and in the conservative two-state Markov model.

In one-way sensitivity analyses, the modeled cost of ECT was the most influential parameter: TAO-TMS lost dominance when the ECT treatment cost fell below US$539. Probabilistic simulations estimated a >85% probability that TAO-TMS was cost-effective at willingness-to-pay thresholds aligned with Singapore gross domestic product (GDP) per capita.

These findings are broadly consistent with previous economic evaluations of TMS and ECT in Singapore (Zhao et al., 2018; Teng et al., 2021), North America (Kozel et al., 2004; Fitzgibbon et al., 2020) and Europe (Zemplényi et al., 2022), and extend this literature to accelerated, connectivity-guided TMS.

Because ECT eligibility was not assessed, we cannot determine whether all TAO-TMS participants would have been ECT candidates. The analysis therefore combined outcomes from this single-arm study with literature-derived ECT parameters from nonpsychotic TRD populations. Although 38% of our participants had previously received scalp-based TMS, ECT populations may include a greater proportion of patients referred after TMS nonresponse. Prospective comparisons among patients eligible for both treatments, ideally randomized or otherwise carefully balanced and with longer follow-up, are needed to refine these estimates and determine whether the projected benefits persist.

### Limitations

Our small sample size offered limited statistical power for subgroup and neuroimaging analyses and increased uncertainty in the health economic estimates. Another key limitation is the lack of a placebo arm or anatomical/scalp-based target as an active control. We are addressing this question in an ongoing double-blind trial directly comparing TAO-TMS with BeamF3 targeting in a naturalistic TRD population with complex psychiatric comorbidities (NCT06385223).

The durability of clinical benefit also warrants consideration. Although MADRS scores remained significantly below baseline at both 1- and 3-month follow-up, the magnitude of improvement was greatest immediately after treatment and attenuated over time. Future studies should evaluate the longer-term durability of benefit and the potential role of maintenance treatment strategies.

Although the relative network contributions varied across participants, TAO-TMS E-field hotspots were generally concentrated within a limited set of networks (Figure 4C). This limited between-participant variation precluded a robust assessment of the relationship between network engagement and clinical outcome. Studies designed to compare targets in different networks are needed to address this question.

Several studies have proposed incorporating E-field modeling to improve personalized targeting (Weise et al., 2020; Lynch et al., 2022; Deng et al., 2023; Elbau et al., 2023; Lueckel et al., 2023; Dannhauer et al., 2024; Sun et al., 2025). The current TAO-TMS trial did not include E-field modeling prospectively. In post hoc exploratory analyses, optimizing coil orientation was not significantly associated with clinical improvement. These analyses were limited by our small sample size, and larger randomized studies are needed to determine whether integrating E-field modeling improves targeting or clinical outcomes.

### Conclusions

This open-label, single-arm proof-of-concept study supports the feasibility and tolerability of connectivity-guided accelerated iTBS in a naturalistic Asian TRD population that included psychiatric comorbidities commonly excluded from previous accelerated iTBS trials. These promising but preliminary clinical outcomes warrant evaluation in larger randomized controlled trials.

## Data Availability

All data produced in the present study are available upon reasonable request to the authors

## Author Contributions

RK, XWT, AX, LQRO, SEG, JJL, TWKT, RSYT, JZJK, HKGS, and PCT contributed to data collection at IMH and NUS. RK, XWT, JC, RSYT, JZJK, JHC, and LS analyzed the data. RK, BTTY and PCT wrote the first draft of the manuscript. All authors edited the manuscript.

## Acknowledgements

The authors would like to acknowledge the support of the Temasek Foundation Climate and Livability Board, the team from the Neurostimulation Service, Institute of Mental Health (Singapore) and our patients and families. Our research is supported by the NUS Yong Loo Lin School of Medicine (NUHSRO/2020/124/TMR/LOA), the Singapore National Medical Research Council (NMRC) LCG (OFLCG19May-0035), NMRC CTG-IIT (CTGIIT23jan-0001), NMRC OF-IRG (OFIRG24jan-0006; OFIRG24jul-0049), NMRC STaR (STaR20nov-0003), Singapore Ministry of Health (MOH) Centre Grant (CG21APR1009), the Temasek Foundation (TF2223-IMH-01), the United States National Institutes of Health (R01MH133334 & 2R01MH120080), and the Singapore National Research Foundation (NRF) Investigatorship (NRFI10-2024-0014). Any opinions, findings and conclusions or recommendations expressed in this material are those of the authors and do not reflect the views of the funders.

## Statement of Ethics

The study was approved by the National Healthcare Group Domain Specific Review Board (DSRB 2023/00397). Written informed consent was obtained from participants (or their parent/legal guardian/next of kin) to participate in the study.

## Supplementary Material

This supplementary material is divided into Supplementary Methods and Supplementary Results to complement the Methods and Results sections in the main text, respectively.

## Supplementary Methods

### MRI acquisition

All MRI scans were acquired at the Centre for Translational Magnetic Resonance Research (TMR), National University of Singapore, on a 3T Siemens Prisma Fit scanner. Each participant underwent a T1-weighted structural scan and two 10-minute resting-state fMRI (rs-fMRI) runs prior to treatment. The structural data were obtained with a T1w MPRAGE sequence using the following acquisition parameters: TR = 2200 ms; TE = 2.45 ms; FOV = 256 × 232 mm; voxel size = 1.0 × 1.0 × 1.0 mm. The rs-fMRI data were obtained with the following acquisition parameters: TR = 1000 ms; TE = 12, 29.75, 47.5 ms; multiband factor 4; flip angle = 50°; FOV = 240 × 240 mm; voxel size = 3.0 × 3.0 × 3.0 mm. During the 10-minute rs-fMRI scans, participants were instructed to keep their eyes open, maintain fixation on a central cross, let their minds wander naturally, and avoid repetitive thoughts. The scalp-based BeamF3 (Beam et al., 2009) stimulation location was also identified and marked on the scalp using a vitamin E capsule. Participants completed a second MRI session on average 5.0 ± 1.9 days after treatment, consisting of one T1-weighted structural scan and two resting-state fMRI runs acquired using the same imaging protocol.

### fMRI preprocessing

T1-weighted MRI data was processed using FreeSurfer (Fischl, 2012). Resting-state fMRI data were preprocessed using the Computational Brain Imaging Group (CBIG) fMRI preprocessing pipeline (Kong et al., 2019; Li et al., 2019; Ooi et al., 2025). Preprocessing included the following steps. (1) Removal of the first 4 frames; (2) Slice time correction; (3) Motion correction and outlier detection: frames with FD > 0.2 mm or DVARS > 50 were flagged as censored frames. 1 frame before and 2 frames after these volumes were flagged as censored frames. Uncensored segments of data lasting fewer than five contiguous frames were also labeled as censored frames. Runs with over half of the frames censored were removed. After censoring, some runs contained less than 10 min of usable data, but in the Results section, they are still referred to as 10-min runs. (4) Correcting for susceptibility-induced spatial distortion; (5) Multi-echo denoising (DuPre et al., 2021); (6) Alignment with the structural image using boundary-based registration (Greve and Fischl, 2009); (7) Global, white matter and ventricular signals, 6 motion parameters, and their temporal derivatives were regressed from the functional data. Regression coefficients were estimated from uncensored data; (8) Censored frames were interpolated with the Lomb-Scargle periodogram (Power et al., 2014); (9) Bandpass filtering (0.009 Hz – 0.08 Hz) was applied to the data; (10) The data was then projected onto FreeSurfer fsaverage6 surface space and smoothed using a 6-mm full-width at half maximum kernel; (11) Lastly, preprocessed data in native fMRI volumetric space was smoothed using a 4-mm full-width at half maximum kernel. Alignment between T1 and MNI152 was also computed using ANTs registration (Avants et al., 2011).

### Exploratory health economic analysis

An exploratory model-based cost-effectiveness analysis compared TAO-TMS with electroconvulsive therapy (ECT) over a one-year time horizon. We used a hybrid decision tree–Markov model with the same structure for both treatment arms (Figure S1). The decision tree represented acute treatment outcomes at the fixed assessment immediately after treatment. Patients were classified as responders (≥ 50% reduction in MADRS), partial responders (≥ 25% but <50% reduction), or nonresponders (<25% reduction). All three branches, including nonresponse, were retained in the model. At 1 month after treatment, patients from each acute-outcome category entered the Markov model and were assigned to one of three health states based on their MADRS score at that assessment: remission (MADRS <11), partial relapse (MADRS 11-20), or relapse (MADRS >20). Thus, the decision tree characterized percentage improvement immediately after treatment, whereas the Markov states characterized absolute symptom severity during follow-up. The Markov model used monthly cycles through month 12. During each cycle, patients could remain in their current health state or transition to another state.

Treatment-specific probabilities were assigned to this common model structure. For TAO-TMS, data from the present trial determined the probabilities of the acute-outcome categories and the probabilities of moving from each acute-outcome category immediately after treatment to each Markov health state at 1 month after treatment. Subsequent transition probabilities among the health states were estimated using changes observed between the 1-month and 3-month assessments. Analyses used patients with the observations required to estimate each probability; missing outcomes were not imputed. Because the 1-month and 3-month assessments were separated by two months, the observed two-month transition probabilities were converted into monthly probabilities. The resulting monthly probabilities were applied through month 12. This extrapolation assumes that transition probabilities observed between months 1 and 3 remained constant during subsequent follow-up.

For ECT, acute-outcome and follow-up probabilities were obtained from published economic evaluations of nonpsychotic treatment-resistant depression populations (Fitzgibbon et al., 2020; Zhao et al., 2018). Because TAO-TMS outcomes were unavailable beyond 3 months and the published ECT inputs did not include recovery from relapse, relapse was treated as an absorbing state in both arms: once patients entered relapse, they remained in that state for the remainder of the modeled period. All clinical probabilities and their sources are reported in Table S6.

Health-state utility values were derived from EQ-5D data collected in the present trial. These utility values were used to estimate quality-adjusted life-years (QALYs). Analyses were conducted from healthcare system and societal perspectives. The healthcare system perspective included treatment, imaging, outpatient visits, medication, laboratory testing, and adverse-event costs. The societal perspective additionally included transportation and productivity losses. Cost inputs were obtained from published Singapore Ministry of Health rates, Singapore Agency for Care Effectiveness resource sheets, institutional cost data, and data from the present trial. All model inputs and their sources are summarized in Table S6.

All costs were expressed in 2025 Singapore dollars. Results were also converted to US dollars using a fixed exchange rate of US$1 = S$1.29. Costs and QALYs were not discounted because the model horizon did not exceed one year. The primary outcomes from the cost-effectiveness analysis (CEA) model were costs, QALYs gained, and the incremental cost-effectiveness ratio (ICER). A willingness-to-pay threshold of S$100,000 (US$77,519) was used.

One-way and probabilistic sensitivity analyses were performed to assess the robustness of the base-case results to parameter uncertainty across different willingness-to-pay thresholds. The probabilistic sensitivity analysis used 10,000 Monte Carlo simulations. These simulations were used to construct cost-effectiveness acceptability curves showing the probability that TAO-TMS was cost-effective relative to ECT across a range of willingness-to-pay thresholds.

Scenario analyses examined 3-month and 6-month time horizons to reduce reliance on extrapolation beyond the observed TAO-TMS follow-up. An additional conservative scenario analysis used a two-state Markov model in which partial relapse and relapse were combined into a single state, leaving remission and relapse as the two health states. Results of the base-case and scenario analyses are reported in Tables S7–S9.

Data preparation and health economic analyses were performed using RStudio Desktop Pro version 2024.04.1 (Posit Software, PBC, Boston, MA). Decision-analytic modeling was conducted using TreeAge Pro 2023 R2 (TreeAge Software, LLC, Williamstown, MA).

## Supplementary Results

### Exploratory health economic outcomes

An exploratory hybrid decision tree and Markov cohort model (Figure S1) was used to assess the costs and effectiveness of TAO-TMS and ECT, utilizing data collected from the study and relevant literature (Table S6). From the perspective of the healthcare system, the base-case model estimated costs that were US$8,214 lower for TAO-TMS than for ECT, with costs amounting to US$35,017 for TAO-TMS versus US$43,230 for ECT. Additionally, TAO-TMS resulted in a higher QALY of 0.67, compared to 0.65 for ECT. When viewed from a societal perspective, estimated costs were US$14,894 lower for TAO-TMS than for ECT. Under the base-case assumptions, TAO-TMS was dominant from both perspectives, with lower estimated costs and similar QALYs (Table S7).

Tornado diagrams were employed to illustrate the outcomes of the one-way sensitivity analyses (Figure S9). From the healthcare system perspective, the modeled cost of ECT was the most influential input parameter. Specifically, if the cost of ECT treatment were to decrease to below US$539 (S$695), the ICER would exceed the WTP threshold, thereby altering the conclusion regarding the cost-effectiveness of TAO-TMS. From the societal perspective, the conclusion favoring TAO-TMS remained unchanged across the one-way sensitivity ranges.

In the probabilistic sensitivity analyses, we conducted 10,000 Monte Carlo simulations, varying the input parameters simultaneously. The results were presented in cost-effectiveness acceptability curves (CEACs), which illustrate the probabilities of each intervention arm being cost-effective across a range of WTP thresholds. The CEACs indicated a high model-estimated probability that TAO-TMS was cost-effective across the evaluated willingness-to-pay thresholds from both perspectives (Figure 7).

In scenarios involving shorter time horizons, the cost savings and QALY gains associated with TAO-TMS were diminished across all scenarios (Table S8), but TAO-TMS remained the preferred option under the modeled assumptions. In a conservative scenario that considered only two treatment responses and two Markov health states (Figure S10), TAO-TMS still demonstrated cost savings relative to ECT (Table S9). However, the QALY gains were found to be comparable between the two treatments, with ECT yielding 0.01 QALY gain compared to TAO-TMS. In this conservative scenario, the ICER for ECT relative to TAO-TMS was US$1,221,700 per QALY from the healthcare system perspective and US$506,450 per QALY from the societal perspective. Both ICERs substantially exceeded the willingness-to-pay threshold, indicating that the additional QALYs associated with ECT were not cost-effective under the assumptions of this scenario.

---

**Table S1.**
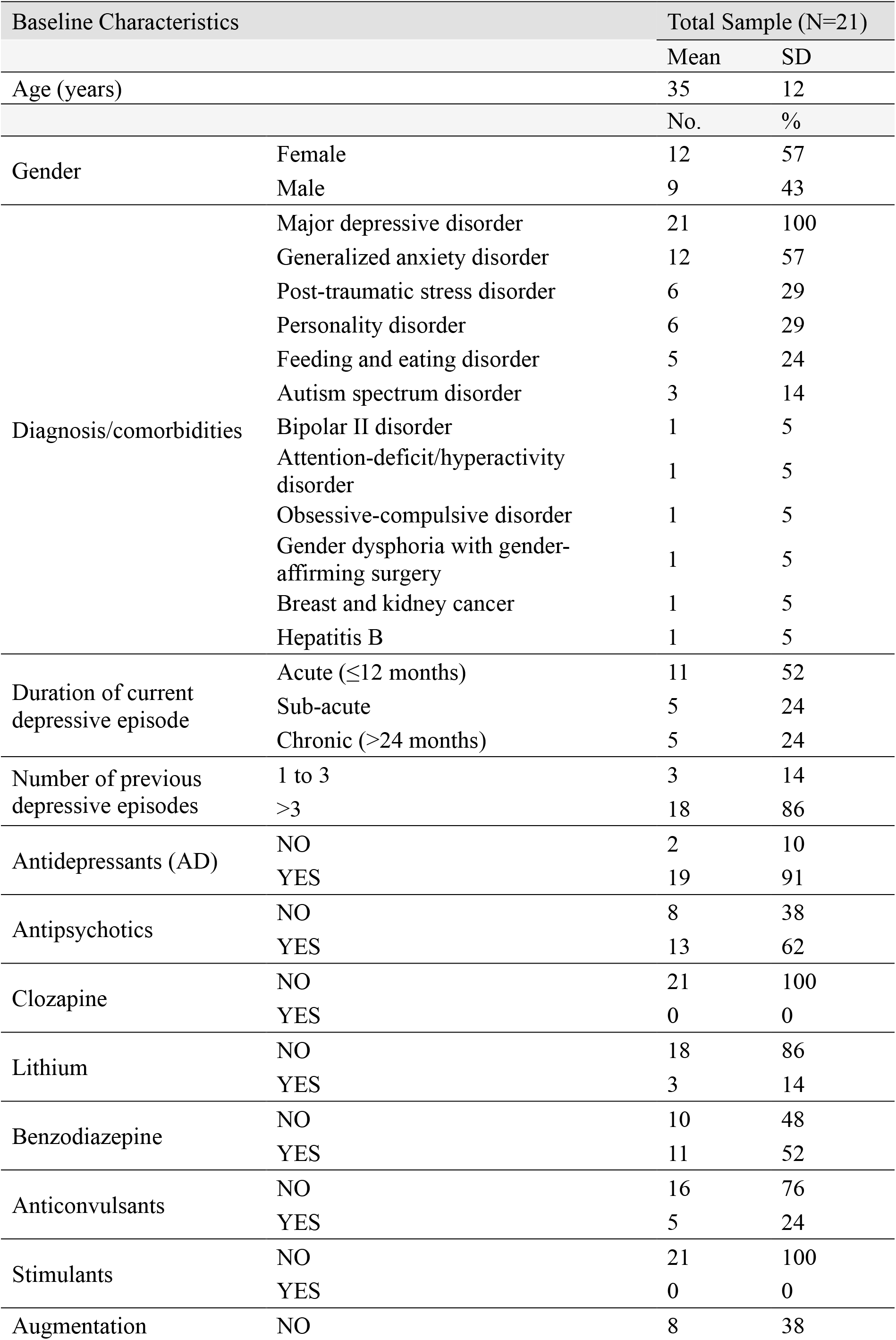
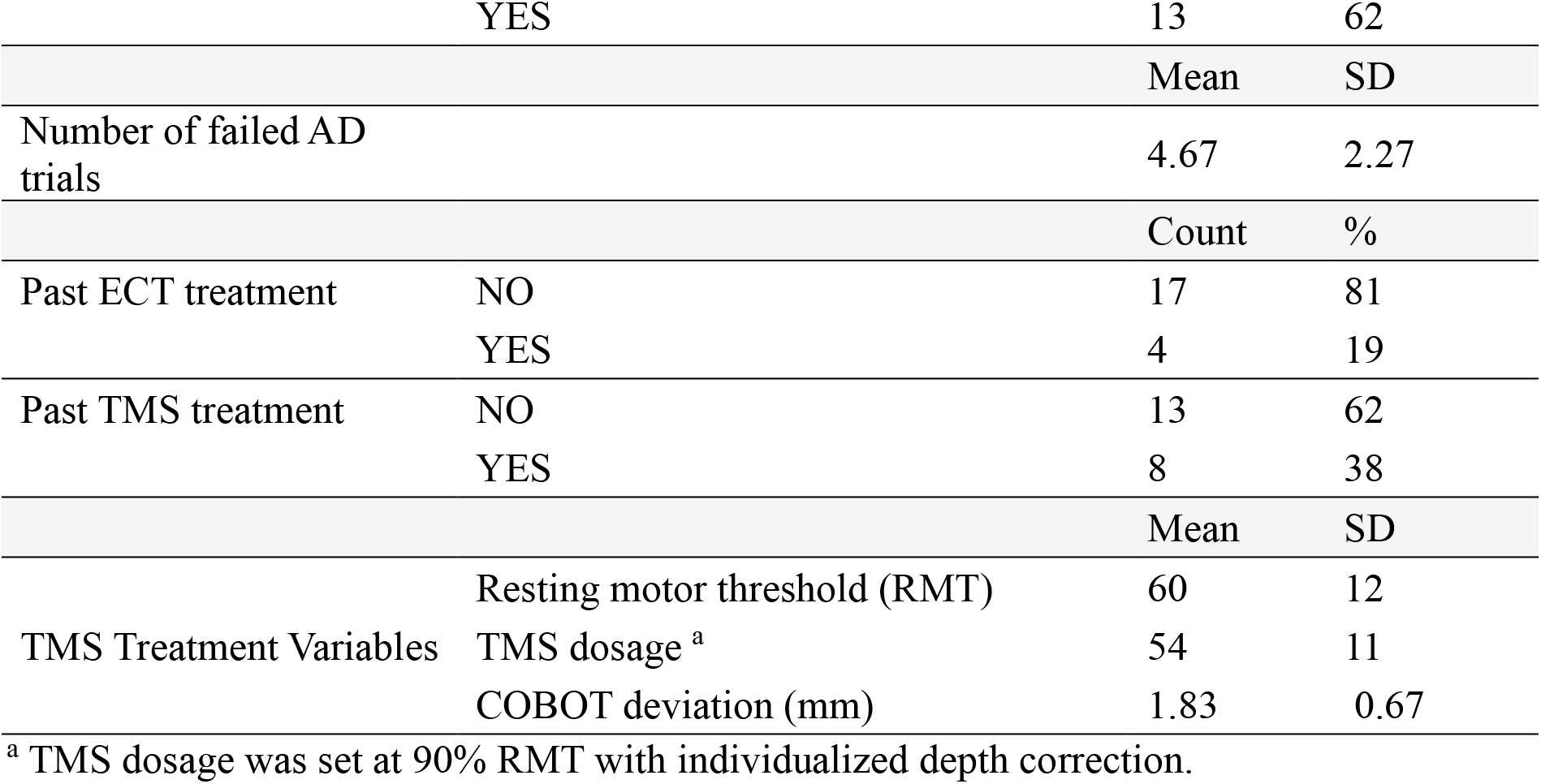
Patient socio-demographics, clinical characteristics, and TMS parameters.

| Baseline Characteristics |  | Total Sample (N=21) |  |
|---|---|---|---|
|  |  | Mean | SD |
| Age (years) |  | 35 | 12 |
|  |  | No. | % |
| Gender | Female | 12 | 57 |
|  | Male | 9 | 43 |
| Diagnosis/comorbidities | Major depressive disorder | 21 | 100 |
|  | Generalized anxiety disorder | 12 | 57 |
|  | Post-traumatic stress disorder | 6 | 29 |
|  | Personality disorder | 6 | 29 |
|  | Feeding and eating disorder | 5 | 24 |
|  | Autism spectrum disorder | 3 | 14 |
|  | Bipolar II disorder | 1 | 5 |
|  | Attention-deficit/hyperactivity disorder | 1 | 5 |
|  | Obsessive-compulsive disorder | 1 | 5 |
|  | Gender dysphoria with gender-affirming surgery | 1 | 5 |
|  | Breast and kidney cancer | 1 | 5 |
|  | Hepatitis B | 1 | 5 |
| Duration of current depressive episode | Acute (≤12 months) | 11 | 52 |
|  | Sub-acute | 5 | 24 |
|  | Chronic (>24 months) | 5 | 24 |
| Number of previous depressive episodes | 1 to 3 | 3 | 14 |
|  | >3 | 18 | 86 |
| Antidepressants (AD) | NO | 2 | 10 |
|  | YES | 19 | 91 |
| Antipsychotics | NO | 8 | 38 |
|  | YES | 13 | 62 |
| Clozapine | NO | 21 | 100 |
|  | YES | 0 | 0 |
| Lithium | NO | 18 | 86 |
|  | YES | 3 | 14 |
| Benzodiazepine | NO | 10 | 48 |
|  | YES | 11 | 52 |
| Anticonvulsants | NO | 16 | 76 |
|  | YES | 5 | 24 |
| Stimulants | NO | 21 | 100 |
|  | YES | 0 | 0 |
| Augmentation | NO | 8 | 38 |
|  | YES | 13 | 62 |
|  |  | Mean | SD |
| Number of failed AD trials |  | 4.67 | 2.27 |
|  |  | Count | % |
| Past ECT treatment | NO | 17 | 81 |
|  | YES | 4 | 19 |
| Past TMS treatment | NO | 13 | 62 |
|  | YES | 8 | 38 |
|  |  | Mean | SD |
| TMS Treatment Variables | Resting motor threshold (RMT) | 60 | 12 |
|  | TMS dosage<sup>a</sup> | 54 | 11 |
|  | COBOT deviation (mm) | 1.83 | 0.67 |

<sup>a</sup> TMS dosage was set at 90% RMT with individualized depth correction.

**Table S2.** Adverse events reported during TAO-TMS treatment.

| Adverse events | Total Sample (N=21) |  |
|---|---|---|
|  | No. | % |
| Any reported adverse event | 10 | 47.6 |
| No reported adverse event | 11 | 52.4 |
| Treatment-site discomfort or headache during stimulation | 5 | 23.8 |
| Insomnia | 4 | 19.0 |
| Anxiety | 3 | 14.3 |
| Dizziness | 2 | 9.5 |
| Neck or back discomfort | 2 | 9.5 |
| Fatigue | 1 | 4.8 |
| Nausea | 1 | 4.8 |
| Skin irritation at the treatment site | 1 | 4.8 |
| Post-traumatic stress disorder (led to discontinuation) | 1 | 4.8 |

**Table S3.** Clinical assessment scores at the respective timepoints of TAO-TMS^a^.

| Assessment scores |  |  |  |  |  |  |  |  |
|---|---|---|---|---|---|---|---|---|
|  | Baseline |  | Post-treatment |  | 1-month FU |  | 3-month FU |  |
|  | (Mean ± SD) | N | (Mean ± SD) | N | (Mean ± SD) | N | (Mean ± SD) | N |
| MADRS | 28.33 ± 5.14 | 21 | 12.10 ± 5.56 | 20 | 18.06 ± 10.12 | 18 | 20.13 ± 7.71 | 16 |
| QIDS-SR16 | 20.67 ± 6.45 |  | 11.30 ± 6.61 |  | 14.44 ± 8.13 |  | 16.44 ± 8.33 |  |
| GAD-7 | 10.86 ± 5.26 |  | 6.90 ± 5.04 |  | 8.94 ± 5.58 |  | 10.38 ± 3.74 |  |
| Q-LES-Q-SF | 39.00 ± 9.41 |  | 49.10 ± 9.77 |  | 46.11 ± 16.48 |  | 44.69 ± 11.02 |  |
| EQ-5D-3L utility score | 0.39 ± 0.33 |  | 0.60 ± 0.28 |  | 0.51 ± 0.40 |  | 0.41 ± 0.45 |  |
| · Visual analogue scale | 42.62 ± 16.50 |  | 59.65 ± 17.54 |  | 57.78 ± 24.09 |  | 49.69 ± 16.78 |  |
| SDS total score | 17.86 ± 6.46 |  | 14.30 ± 7.66 |  | 11.71 ± 9.48 |  | 13.63 ± 6.35 |  |
| · Days lost | 3.61 ± 2.89 |  | 1.80 ± 2.46 |  | 1.33 ± 1.99 |  | 2.54 ± 2.99 |  |
| · Days underproductive | 5.12 ± 2.20 |  | 2.92 ± 2.50 |  | 3.53 ± 2.90 |  | 3.23 ± 2.80 |  |
| MoCA | 28.52 ± 1.47 |  | 28.00 ± 1.65 |  | 27.59 ± 2.32 |  | 28.73 ± 1.22 |  |
| MADRS-Q10 | 2.52 ± 1.33 |  | 1.25 ± 0.79 |  | 1.78 ± 1.67 |  | 2.25 ± 1.39 |  |
|  | Rate (%) | N | Rate (%) | N | Rate (%) | N | Rate (%) | N |
| Response<sup>a</sup> | - | - | 65 | 13 | 50 | 9 | 31.25 | 5 |
| Remission<sup>b</sup> | - | - | 30 | 6 | 27.78 | 5 | 6.25 | 1 |

Abbreviations: MADRS, Montgomery-Asberg Depression Rating Scale; QIDS-SR16, The Quick Inventory of Depressive Symptomatology (16-Item) (Self-Report); GAD-7, 7-item Generalized Anxiety Disorder scale; Q-LES-Q-SF, Quality of Life Enjoyment and Satisfaction Questionnaire-Short form; EQ-5D-3L, EuroQol 5-dimensional questionnaire with 3 levels; SDS, Sheehan Disability Scale; MoCA, the Montreal Cognitive Assessment; FU: follow-up
<sup>a</sup> Response was defined as a ≥50% reduction in total MADRS score from baseline.
<sup>b</sup> Remission was defined as a total MADRS score of <11.

**Table S4.** Pairwise comparisons between baseline, post-treatment and follow-up assessments for all clinical outcome measures based on linear mixed-effects modeling.

| Measure | Comparisons | Mean difference | SE | df | *p* | 95% CI |  |
|---|---|---|---|---|---|---|---|
|  |  |  |  |  |  | Lower | Upper |
| MADRS | Baseline vs Post-treatment | 16.238* | 1.948 | 51.586 | <0.001 | 10.893 | 21.583 |
|  | Baseline vs 1-month FU | 10.108* | 2.013 | 52.522 | <0.001 | 4.589 | 15.627 |
|  | Baseline vs 3-month FU | 7.975* | 2.090 | 53.331 | 0.002 | 2.248 | 13.702 |
|  | Post-treatment vs 1-month FU | -6.130* | 2.029 | 51.675 | 0.023 | -11.698 | -0.562 |
|  | Post-treatment vs 3-month FU | -8.263* | 2.106 | 52.532 | 0.002 | -14.036 | -2.489 |
|  | 1-month FU vs 3-month FU | -2.133 | 2.158 | 52.513 | 1.000 | -8.049 | 3.783 |
| QIDS-SR16 | Baseline vs Post-treatment | 9.433* | 1.640 | 58.054 | <0.001 | 4.952 | 13.915 |
|  | Baseline vs 1-month FU | 6.177* | 1.699 | 58.657 | 0.004 | 1.538 | 10.816 |
|  | Baseline vs 3-month FU | 4.349 | 1.767 | 59.107 | 0.101 | -0.475 | 9.174 |
|  | Post-treatment vs 1-month FU | -3.256 | 1.708 | 57.821 | 0.370 | -7.924 | 1.411 |
|  | Post-treatment vs 3-month FU | -5.084* | 1.776 | 58.329 | 0.035 | -9.935 | -0.232 |
|  | 1-month FU vs 3-month FU | -1.828 | 1.820 | 58.267 | 1.000 | -6.798 | 3.143 |
| GAD-7 | Baseline vs Post-treatment | 4.055* | 0.877 | 51.867 | <0.001 | 1.650 | 6.461 |
|  | Baseline vs 1-month FU | 1.761 | 0.909 | 52.161 | 0.350 | -0.733 | 4.255 |
|  | Baseline vs 3-month FU | 1.308 | 0.947 | 52.358 | 1.000 | -1.289 | 3.904 |
|  | Post-treatment vs 1-month FU | -2.295 | 0.913 | 51.605 | 0.090 | -4.799 | 0.209 |
|  | Post-treatment vs 3-month FU | -2.748* | 0.950 | 51.844 | 0.033 | -5.354 | -0.142 |
|  | 1-month FU vs 3-month FU | -0.453 | 0.973 | 51.798 | 1.000 | -3.122 | 2.216 |
| Q-LES-Q-SF | Baseline vs Post-treatment | -10.088* | 2.626 | 51.720 | 0.002 | -17.292 | -2.884 |
|  | Baseline vs 1-month FU | -8.800* | 2.717 | 52.456 | 0.013 | -16.251 | -1.350 |
|  | Baseline vs 3-month FU | -4.170 | 2.803 | 52.206 | 0.857 | -11.857 | 3.518 |
|  | Post-treatment vs 1-month FU | 1.288 | 2.692 | 51.093 | 1.000 | -6.100 | 8.676 |
|  | Post-treatment vs 3-month FU | 5.919 | 2.799 | 51.798 | 0.236 | -1.760 | 13.597 |
|  | 1-month FU vs 3-month FU | 4.631 | 2.868 | 51.790 | 0.675 | -3.238 | 12.499 |
| EQ-5D-3L | Baseline vs Post-treatment | -0.212 | 0.088 | 52.631 | 0.116 | -0.452 | 0.029 |
|  | Baseline vs 1-month FU | -0.117 | 0.091 | 53.292 | 1.000 | -0.366 | 0.131 |
|  | Baseline vs 3-month FU | 1.707e-5 | 0.094 | 53.810 | 1.000 | -0.258 | 0.258 |
|  | Post-treatment vs 1-month FU | 0.094 | 0.091 | 52.507 | 1.000 | -0.156 | 0.345 |
|  | Post-treatment vs 3-month FU | 0.212 | 0.095 | 53.077 | 0.180 | -0.048 | 0.472 |
|  | 1-month FU vs 3-month FU | 0.117 | 0.097 | 53.028 | 1.000 | -0.149 | 0.384 |
| SDS | Baseline vs Post-treatment | 4.795* | 1.374 | 51.950 | 0.006 | 1.027 | 8.564 |
|  | Baseline vs 1-month FU | 5.971* | 1.424 | 52.310 | <0.001 | 2.066 | 9.877 |
|  | Baseline vs 3-month FU | 3.322 | 1.482 | 52.560 | 0.176 | -0.742 | 7.387 |
|  | Post-treatment vs 1-month FU | 1.176 | 1.430 | 51.687 | 1.000 | -2.747 | 5.099 |
|  | Post-treatment vs 3-month FU | -1.473 | 1.488 | 51.982 | 1.000 | -5.555 | 2.608 |
|  | 1-month FU vs 3-month FU | -2.649 | 1.524 | 51.931 | 0.529 | -6.830 | 1.532 |
| MoCA | Baseline vs Post-treatment | 0.496 | 0.417 | 50.833 | 1.000 | -0.647 | 1.640 |
|  | Baseline vs 1-month FU | 1.105 | 0.439 | 51.994 | 0.090 | -0.100 | 2.310 |
|  | Baseline vs 3-month FU | -0.281 | 0.458 | 52.572 | 1.000 | -1.536 | 0.975 |
|  | Post-treatment vs 1-month FU | 0.609 | 0.442 | 51.242 | 1.000 | -0.605 | 1.823 |
|  | Post-treatment vs 3-month FU | -0.777 | 0.461 | 51.872 | 0.587 | -2.041 | 0.487 |
|  | 1-month FU vs 3-month FU | -1.386* | 0.476 | 51.423 | 0.032 | -2.693 | -0.078 |

MADRS, Montgomery-Asberg Depression Rating Scale; QIDS-SR16, The Quick Inventory of Depressive Symptomatology (16-Item) (Self-Report); GAD-7, 7-item Generalized Anxiety Disorder scale; Q-LES-Q-SF, Quality of Life Enjoyment and Satisfaction Questionnaire-Short form; EQ-5D-3L, EuroQol 5-dimensional questionnaire with 3 levels; SDS, Sheehan Disability Scale; MoCA, the Montreal Cognitive Assessment; FU: follow-up.
\* indicates that the difference is significant at the 0.05 level after Bonferroni correction. The reported p values are Bonferroni-corrected p values.

**Table S5.** Pairwise comparisons of daily QIDS-SR16 and GAD-7 scores across the five treatment days based on linear mixed-effects modeling.

| Measure | Comparisons | Mean difference | SE | df | *p* | 95% CI |  |
|---|---|---|---|---|---|---|---|
|  |  |  |  |  |  | Lower | Upper |
| QIDS-SR16 | Baseline vs Day 1 | -0.350 | 1.242 | 96.615 | 1.000 | -4.089 | 3.390 |
|  | Baseline vs Day 2 | 3.900* | 1.242 | 96.615 | 0.034 | 0.161 | 7.640 |
|  | Baseline vs Day 3 | 5.250* | 1.242 | 96.615 | <0.001 | 1.511 | 8.990 |
|  | Baseline vs Day 4 | 6.400* | 1.242 | 96.615 | <0.001 | 2.661 | 10.140 |
|  | Baseline vs Day 5 | 9.450* | 1.242 | 96.615 | <0.001 | 5.711 | 13.190 |
|  | Day 1 vs Day 2 | 4.250* | 1.248 | 95.451 | 0.015 | 0.493 | 8.007 |
|  | Day 1 vs Day 3 | 5.600* | 1.248 | 95.451 | <0.001 | 1.843 | 9.357 |
|  | Day 1 vs Day 4 | 6.750* | 1.248 | 95.451 | <0.001 | 2.993 | 10.507 |
|  | Day 1 vs Day 5 | 9.800* | 1.248 | 95.451 | <0.001 | 6.043 | 13.557 |
|  | Day 2 vs Day 3 | 1.350 | 1.248 | 95.451 | 1.000 | -2.407 | 5.107 |
|  | Day 2 vs Day 4 | 2.500 | 1.248 | 95.451 | 0.720 | -1.257 | 6.257 |
|  | Day 2 vs Day 5 | 5.550* | 1.248 | 95.451 | <0.001 | 1.793 | 9.307 |
|  | Day 3 vs Day 4 | 1.150 | 1.248 | 95.451 | 1.000 | -2.607 | 4.907 |
|  | Day 3 vs Day 5 | 4.200* | 1.248 | 95.451 | 0.017 | 0.443 | 7.957 |
|  | Day 4 vs Day 5 | 3.050 | 1.248 | 95.451 | 0.245 | -0.707 | 6.807 |
| GAD-7 | Baseline vs Day 1 | 1.326 | 0.610 | 95.830 | 0.484 | -0.511 | 3.163 |
|  | Baseline vs Day 2 | 2.926* | 0.610 | 95.830 | <0.001 | 1.089 | 4.763 |
|  | Baseline vs Day 3 | 3.276* | 0.610 | 95.830 | <0.001 | 1.439 | 5.113 |
|  | Baseline vs Day 4 | 3.776* | 0.610 | 95.830 | <0.001 | 1.939 | 5.613 |
|  | Baseline vs Day 5 | 4.976* | 0.610 | 95.830 | <0.001 | 3.139 | 6.813 |
|  | Day 1 vs Day 2 | 1.600 | 0.611 | 95.169 | 0.155 | -0.241 | 3.441 |
|  | Day 1 vs Day 3 | 1.950* | 0.611 | 95.169 | 0.029 | 0.109 | 3.791 |
|  | Day 1 vs Day 4 | 2.450* | 0.611 | 95.169 | 0.002 | 0.609 | 4.291 |
|  | Day 1 vs Day 5 | 3.650* | 0.611 | 95.169 | <0.001 | 1.809 | 5.491 |
|  | Day 2 vs Day 3 | 0.350 | 0.611 | 95.169 | 1.000 | -1.491 | 2.191 |
|  | Day 2 vs Day 4 | 0.850 | 0.611 | 95.169 | 1.000 | -0.991 | 2.691 |
|  | Day 2 vs Day 5 | 2.050* | 0.611 | 95.169 | 0.017 | 0.209 | 3.891 |
|  | Day 3 vs Day 4 | 0.500 | 0.611 | 95.169 | 1.000 | -1.341 | 2.341 |
|  | Day 3 vs Day 5 | 1.700 | 0.611 | 95.169 | 0.098 | -0.141 | 3.541 |
|  | Day 4 vs Day 5 | 1.200 | 0.611 | 95.169 | 0.789 | -0.641 | 3.041 |

QIDS-SR16, The Quick Inventory of Depressive Symptomatology (16-Item) (Self-Report); GAD-7, 7-item Generalized Anxiety Disorder scale.
\* indicates that the difference is significant at the 0.05 level after Bonferroni correction. The reported p values are Bonferroni-corrected p values.

**Table S6.** Input parameters for the exploratory health economic analysis comparing TAO-TMS and ECT.

| Parameters | Base value (SD) | Low | High | Distribution | Data sources |
|---|---|---|---|---|---|
| Treatment costs (SGD) |  |  |  |  |  |
| fMRI acquisition | 1,158 | 365 | 5,000 | Gamma | ACE resource sheet, hospital database, study data |
| fMRI analysis | 2,000 | 1,000 | 3,000 | Gamma | hospital database |
| aTMS | 150 | 130 | 200 | Gamma | hospital database, study data |
| ECT | 1,700 | 300 | 2,600 | Gamma | MOH, hospital database |
| Health state direct costs (SGD) |  |  |  |  |  |
| Remission |  |  |  |  |  |
| Outpatient visits | 333 (232) | - | - | Gamma | Hospital database, study data |
| Adverse event | 8,988 (8,988) | - | - | Gamma | Hospital database, study data |
| Laboratory tests | 15.6 (15.6) | - | - | Gamma | Hospital database, study data |
| Medications | 965 (2,034) | - | - | Gamma | Hospital database, study data |
| Partial Relapse |  |  |  |  |  |
| Outpatient visits | 325 (223) | - | - | Gamma | Hospital database, study data |
| Adverse event | 10,289.5 (650.75) | - | - | Gamma | Hospital database, study data |
| Laboratory tests | 45.3 (21.2) | - | - | Gamma | Hospital database, study data |
| Medications | 1,254 (3,416) | - | - | Gamma | Hospital database, study data |
| Relapse |  |  |  |  |  |
| Outpatient visits | 488 (321) | - | - | Gamma | Hospital database, study data |
| Adverse event | 11,591 (8,489) | - | - | Gamma | Hospital database, study data |
| Laboratory tests | 88.2 (42.1) | - | - | Gamma | Hospital database, study data |
| Medications | 455 (479) | - | - | Gamma | Hospital database, study data |
| Indirect costs (SGD) |  |  |  |  |  |
| Transport to outpatient visits | 8.13 (6.78) | - | - |  |  |
| Transport to adverse event episode | 16 (16) | - | - |  |  |
| Transport to laboratory test | 3.2 (3.2) | - | - |  |  |
| Median salary | 5,500 (5,500) | - | - |  |  |
| Utility scores |  |  |  |  |  |
| Treatment-resistant depression | 0.524 (0.27) | - | - | Beta | Study data |
| Response | 0.747 (0.184) | - | - | Beta | Study data |
| Partial response | 0.66 (0.22) | - | - | Beta | Study data |
| Remission | 0.684 (0.2) | - | - | Beta | Study data |
| Partial relapse | 0.641 (0.197) | - | - | Beta | Study data |
| Relapse | 0.54 (0.152) | - | - | Beta | Study data |
| Initial decision tree probabilities |  |  |  |  |  |
| Response in TAO-TMS | 0.65 (0.325) | - | - | - | Study data |
| Partial response in TAO-TMS | 0.35 (0.175) | - | - | - | Study data |
| Response in ECT | 0.666 (0.333) | - | - | - | Fitzgibbon et al., 2020 |
| Initial Markov probabilities (at 1 month) |  |  |  |  |  |
| TAO-TMS (conditional probabilities from acute outcome to 1 month health state) |  |  |  |  |  |
| Response to remission | 0.46 | - | - | - | Study data |
| Response to partial relapse | 0.54 | - | - | - | Study data |
| Partial response to remission | 0 | - | - | - | Study data |
| Partial response to partial relapse | 0.71 | - | - | - | Study data |
| No response to remission | 0 | - | - | - | Study data |
| No response to partial relapse | 0 | - | - | - | Study data |
| ECT (initial probabilities at 1 month) |  |  |  |  |  |
| Probability of remission | 0.509 | - | - | - | Fitzgibbon et al., 2020 |
| Probability of partial relapse | 0 | - | - | - | No data |
| Markov transition (monthly transition probabilities) |  |  |  |  |  |
| TAO-TMS |  |  |  |  |  |
| Remission to partial relapse | 0.225 | - | - | - | Study data |
| Remission to relapse | 0.225 | - | - | - | Study data |
| Partial relapse to remission | 0 | - | - | - | Study data |
| Partial relapse to relapse | 0.181 | - | - | - | Study data |
| Relapse to remission | 0 | - | - | - | Study data |
| Relapse to partial relapse | 0 | - | - | - | Study data |
| ECT |  |  |  |  |  |
| Remission to partial relapse | 0 | - | - | - | No data |
| Remission to relapse | 0.34 | - | - | - | Zhao et al., 2018 |
| Partial relapse to remission | 0.7 | - | - | - | Zhao et al., 2018 |
| Partial relapse to relapse | 0.3 | - | - | - | Zhao et al., 2018 |
| Relapse to remission | 0 | - | - | - | No data |
| Relapse to partial relapse | 0 | - | - | - | No data |

SD: Standard deviation. All costs are expressed in 2025 Singapore Dollars (SGD).

**Table S7.** Base-case cost-effectiveness results from health system and societal perspectives.

| Perspectives | Strategies | Costs (SGD) | Incremental cost (SGD) | QALY | Incremental QALY | ICER (SGD/QALY) |
|---|---|---|---|---|---|---|
| Health system | TAO-TMS | 45,172 | - | 0.67 | - | - |
|  | ECT | 55,767 | 10,596 | 0.65 | -0.01 | -722,122 |
| Societal | TAO-TMS | 126,895 | - | 0.67 | - | - |
|  | ECT | 146,109 | 19,213 | 0.65 | -0.01 | -1,309,435 |
| Perspectives | Strategies | Costs (USD) | Incremental cost (USD) | QALY | Incremental QALY | ICER (USD/QALY) |
| Health system | TAO-TMS | 35,017 | - | 0.67 | - | - |
|  | ECT | 43,230 | 8,214 | 0.65 | -0.01 | -559,784 |
| Societal | TAO-TMS | 98,368 | - | 0.67 | - | - |
|  | ECT | 113,263 | 14,894 | 0.65 | -0.01 | -1,015,066 |

TAO-TMS: Tree-based Algorithm for Optimized Transcranial Magnetic Stimulation, ECT: electroconvulsive therapy; QALY: quality-adjusted life-year; ICER: incremental cost-effectiveness ratio. All costs were analyzed in 2025 Singapore Dollars (SGD) and converted to US Dollars (USD) with currency conversion of 1 USD = 1.29 SGD.

**Table S8.** Scenario analyses examining cost-effectiveness of TAO-TMS over shorter time horizons (3-month and 6-month models).

| Scenarios | Strategies | Costs (SGD) | Incremental cost (SGD) | QALY | Incremental QALY | ICER (SGD/QALY) |
|---|---|---|---|---|---|---|
| Health system |  |  |  |  |  |  |
| Time horizon of 3 months | TAO-TMS | 18,686 | - | 0.26 | - | - |
|  | ECT | 28,777 | 10,091 | 0.25 | -0.01 | -1,255,131 |
| Time horizon of 6 months | TAO-TMS | 27,341 | - | 0.39 | - | - |
|  | ECT | 37,662 | 10,321 | 0.38 | -0.01 | -936,854 |
| Societal |  |  |  |  |  |  |
| Time horizon of 3 months | TAO-TMS | 33,918 | - | 0.26 | - | - |
|  | ECT | 47,725 | 13,807 | 0.25 | -0.01 | -1,717,369 |
| Time horizon of 6 months | TAO-TMS | 63,173 | - | 0.39 | - | - |
|  | ECT | 79,487 | 16,314 | 0.38 | -0.01 | -1,480,804 |
| Scenarios | Strategies | Costs (USD) | Incremental cost (USD) | QALY | Incremental QALY | ICER (USD/QALY) |
| Health system |  |  |  |  |  |  |
| Time horizon of 3 months | TAO-TMS | 14,485 | - | 0.26 | - | - |
|  | ECT | 22,308 | 7,822 | 0.25 | -0.01 | -972,970 |
| Time horizon of 6 months | TAO-TMS | 21,194 | - | 0.39 | - | - |
|  | ECT | 29,195 | 8,001 | 0.38 | -0.01 | -726,243 |
| Societal |  |  |  |  |  |  |
| Time horizon of 3 months | TAO-TMS | 26,293 | - | 0.26 | - | - |
|  | ECT | 36,996 | 10,703 | 0.25 | -0.01 | -1,331,294 |
| Time horizon of 6 months | TAO-TMS | 48,971 | - | 0.39 | - | - |
|  | ECT | 61,618 | 12,646 | 0.38 | -0.01 | -1,147,910 |

TAO-TMS: Tree-based Algorithm for Optimized Transcranial Magnetic Stimulation, ECT: electroconvulsive therapy; QALY: quality-adjusted life-year; ICER: incremental cost-effectiveness ratio. All costs were analyzed in 2025 Singapore Dollars (SGD) and converted to US Dollars (USD) with currency conversion of 1 USD = 1.29 SGD.

**Table S9.** Scenario analyses using conservative two-state Markov model structures.

| Perspectives | Strategies | Costs (SGD) | Incremental cost (SGD) | QALY | Incremental QALY | ICER (SGD/QALY) |
|---|---|---|---|---|---|---|
| Health system | TAO-TMS | 45,209 | - | 0.72 | - | - |
|  | ECT | 54,227 | 9,018 | 0.73 | 0.01 | 1,575,993 |
| Societal | TAO-TMS | 122,784 | - | 0.72 | - | - |
|  | ECT | 126,522 | 3,739 | 0.73 | 0.01 | 653,320 |
| Perspectives | Strategies | Costs (USD) | Incremental cost (USD) | QALY | Incremental QALY | ICER (USD/QALY) |
| Health system | TAO-TMS | 35,046 | - | 0.72 | - | - |
|  | ECT | 42,036 | 6,991 | 0.73 | 0.01 | 1,221,700 |
| Societal | TAO-TMS | 95,181 | - | 0.72 | - | - |
|  | ECT | 98,079 | 2,899 | 0.73 | 0.01 | 506,450 |

TAO-TMS: Tree-based Algorithm for Optimized Transcranial Magnetic Stimulation, ECT: electroconvulsive therapy; QALY: quality-adjusted life-year; ICER: incremental cost-effectiveness ratio. All costs were analyzed in 2025 Singapore Dollars (SGD) and converted to US Dollars (USD) with currency conversion of 1 USD = 1.29 SGD.

**Figure S1.**
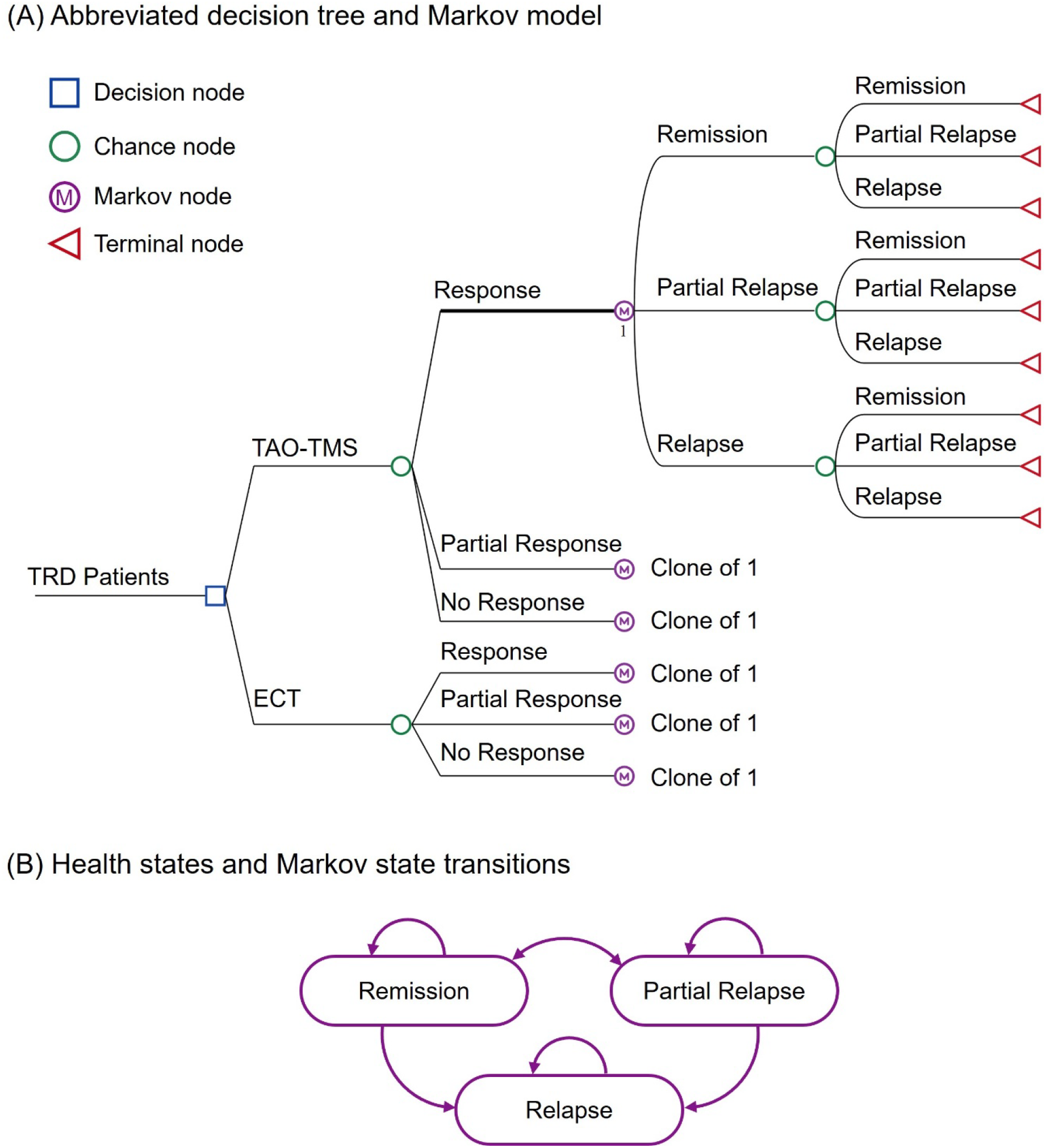
Decision analytical model comparing TAO-TMS and ECT. (A) Abbreviated decision tree and Markov model representing the clinical pathways for patients receiving TAO-TMS or ECT. Initial treatment decisions from the decision tree fed into the Markov model for long-term simulation of outcomes and costs. (B) Health states and Markov state transitions. Arrows indicate possible transitions between health states at each cycle, reflecting remission, partial relapse or relapse under each intervention. Relapse was modeled as an absorbing state, with no transitions from relapse to remission or partial relapse.

**Figure S2.**
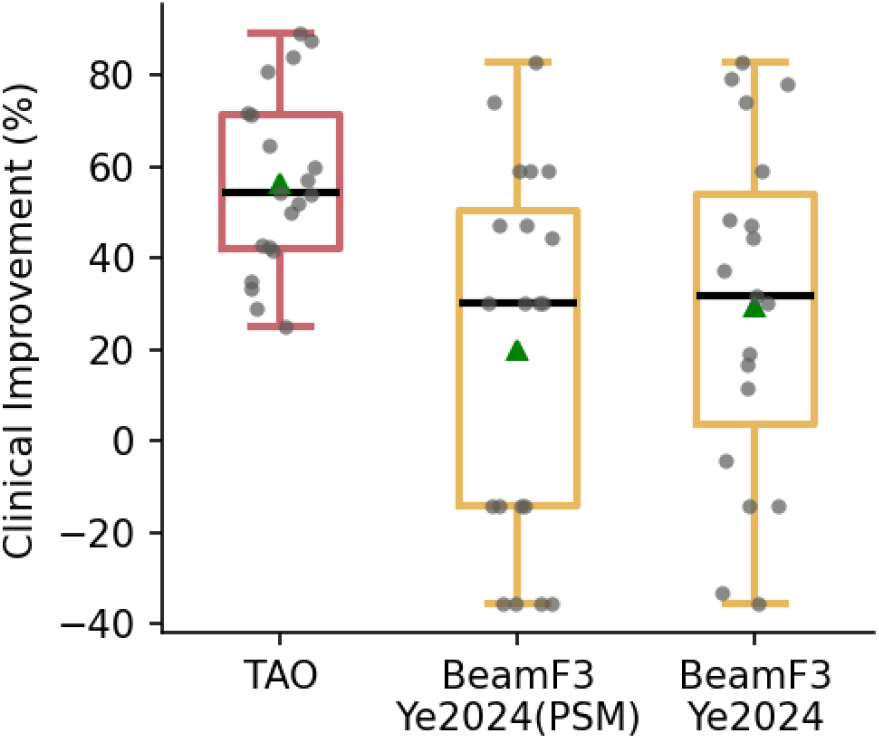
Clinical improvement with TAO-TMS (N = 20) and with non-accelerated BeamF3 TMS in a retrospective sample from the same hospital (N = 23; # sessions = 49 ± 29; Ye et al., 2024). Boxplots show percentage reduction in MADRS scores immediately after treatment. Positive values indicate greater improvement. Females were over-represented in the Ye2024 study, so the middle boxplot “BeamF3 Ye2024 (PSM)” shows the clinical improvement of a subsample of participants that were propensity matched (in terms of baseline MADRS score, number of comorbid psychiatric diagnoses, age, and sex) to participants in the TAO-TMS trial.

**Figure S3.**
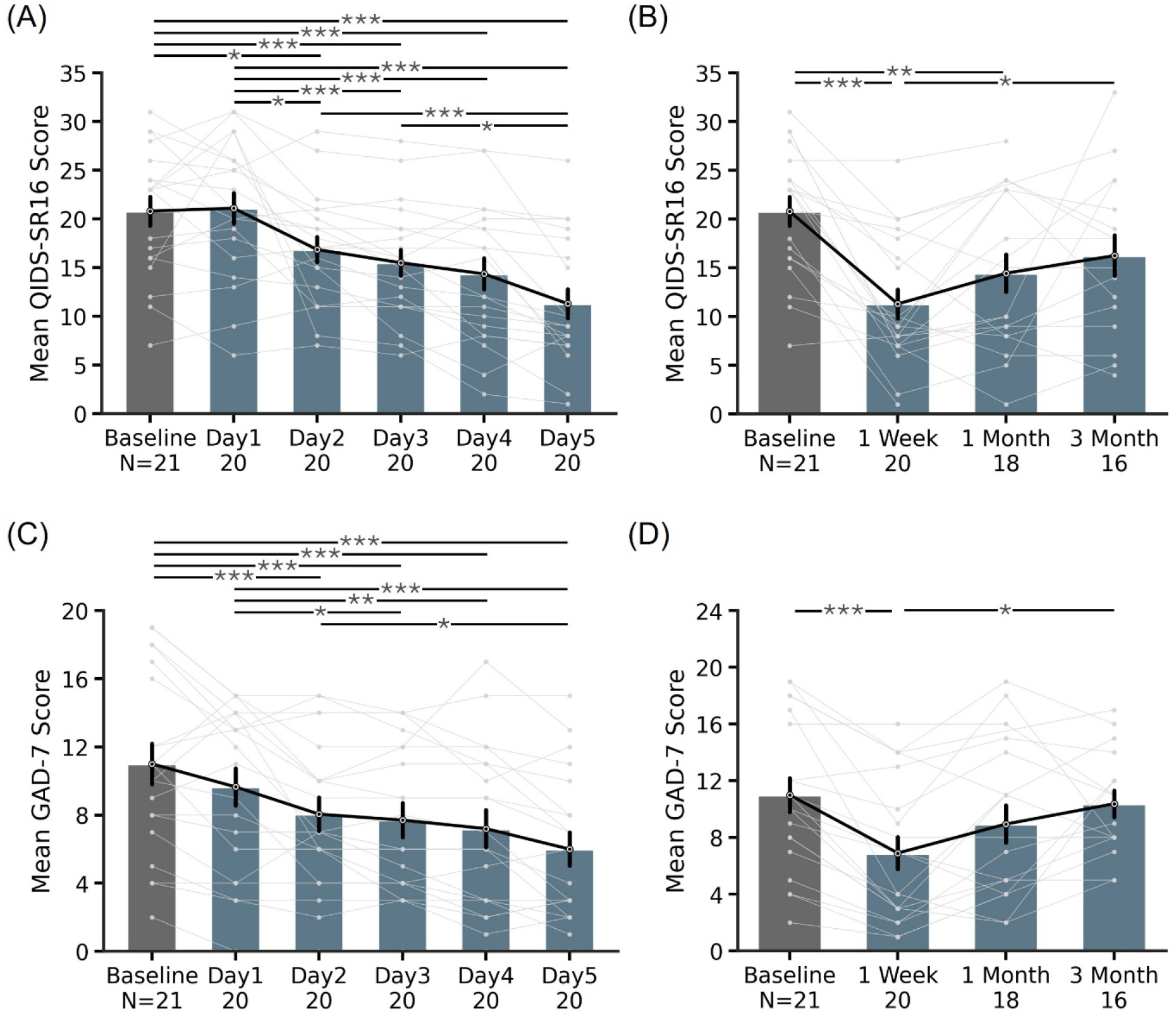
Longitudinal symptom trajectories during and after TAO-TMS treatment. (A) Mean QIDS-SR16 scores from baseline through each day of acute treatment (Day 1 to Day 5). (B) Mean QIDS-SR16 scores at baseline, post-acute treatment (1 week), 1-month, and 3-month follow-up. (C) Mean GAD-7 scores from baseline through each day of acute treatment (Day 1 to Day 5). (D) Mean GAD-7 scores at baseline, post-acute treatment (1 week), 1-month, and 3-month follow-up. Error bars represent standard error. * indicates statistical significance after Bonferroni correction for multiple comparisons. *p<0.05. **p<0.01. ***p<0.001. Bonferroni correction was applied to each measure separately.

**Figure S4.**
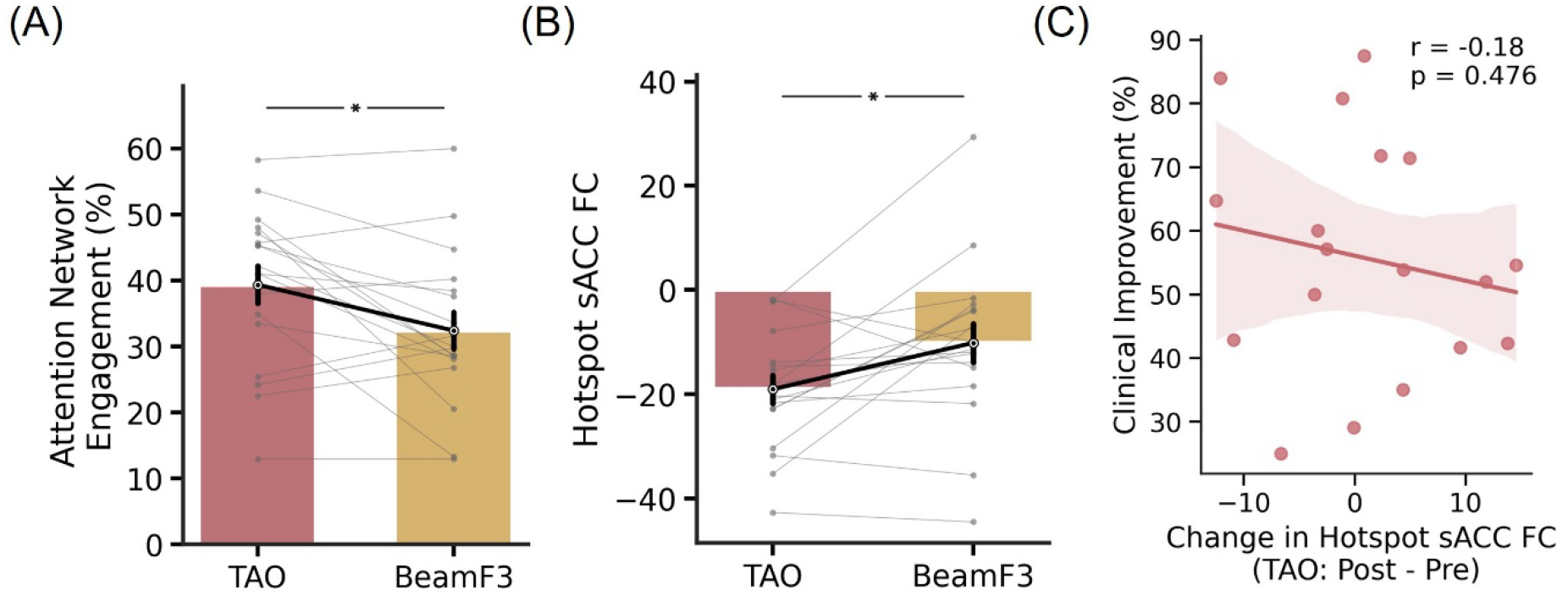
Electric field characteristics of TAO-TMS and BeamF3 targets. (A) Comparison of TAO-TMS and BeamF3 targets in terms of attentional network E-field engagement across 20 participants. Higher values indicate greater engagement of the target networks. (B) Comparison of TAO-TMS targets and BeamF3 targets in terms of hotspot sACC FC across 20 participants. Lower values indicate stronger anticorrelation with sACC. (C) Association between clinical improvement (percentage MADRS reduction) and the change in hotspot sACC FC from pre-treatment to post-treatment. In panels A and B, 20 participants contributed to the plots. In panel C, 18 participants contributed to this plot because one missed the post-treatment scan and one was excluded due to excessive motion.

**Figure S5.**
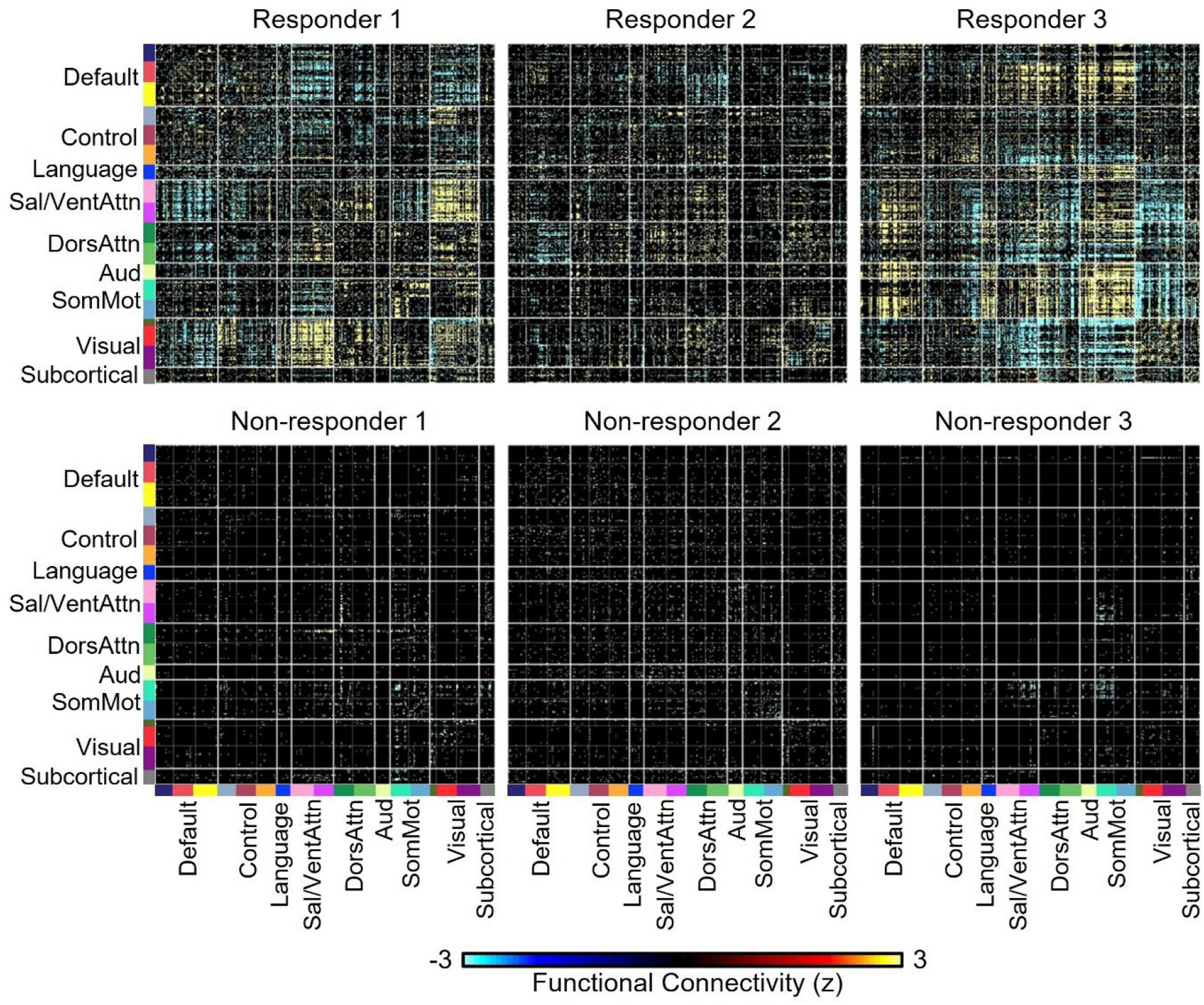
Thresholded functional connectivity (FC) change matrices for six participants. The 419 × 419 matrices show changes in FC from pre-treatment to post-treatment for three clinical responders and three non-responders, with edges surviving false discovery rate (FDR) correction at q < 0.05 highlighted.

**Figure S6.**
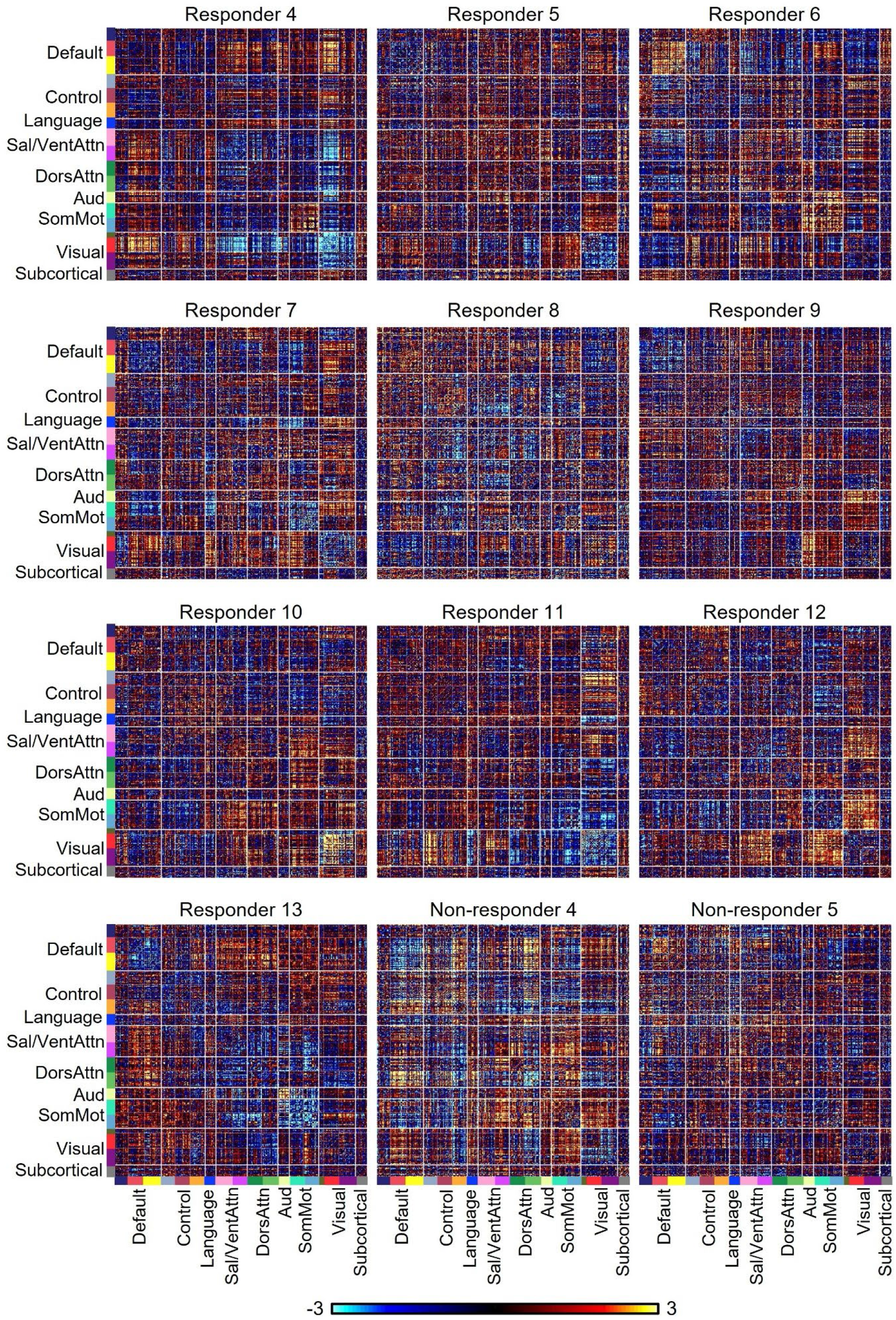
Functional connectivity (FC) change matrices for the remaining participants.

**Figure S7.**
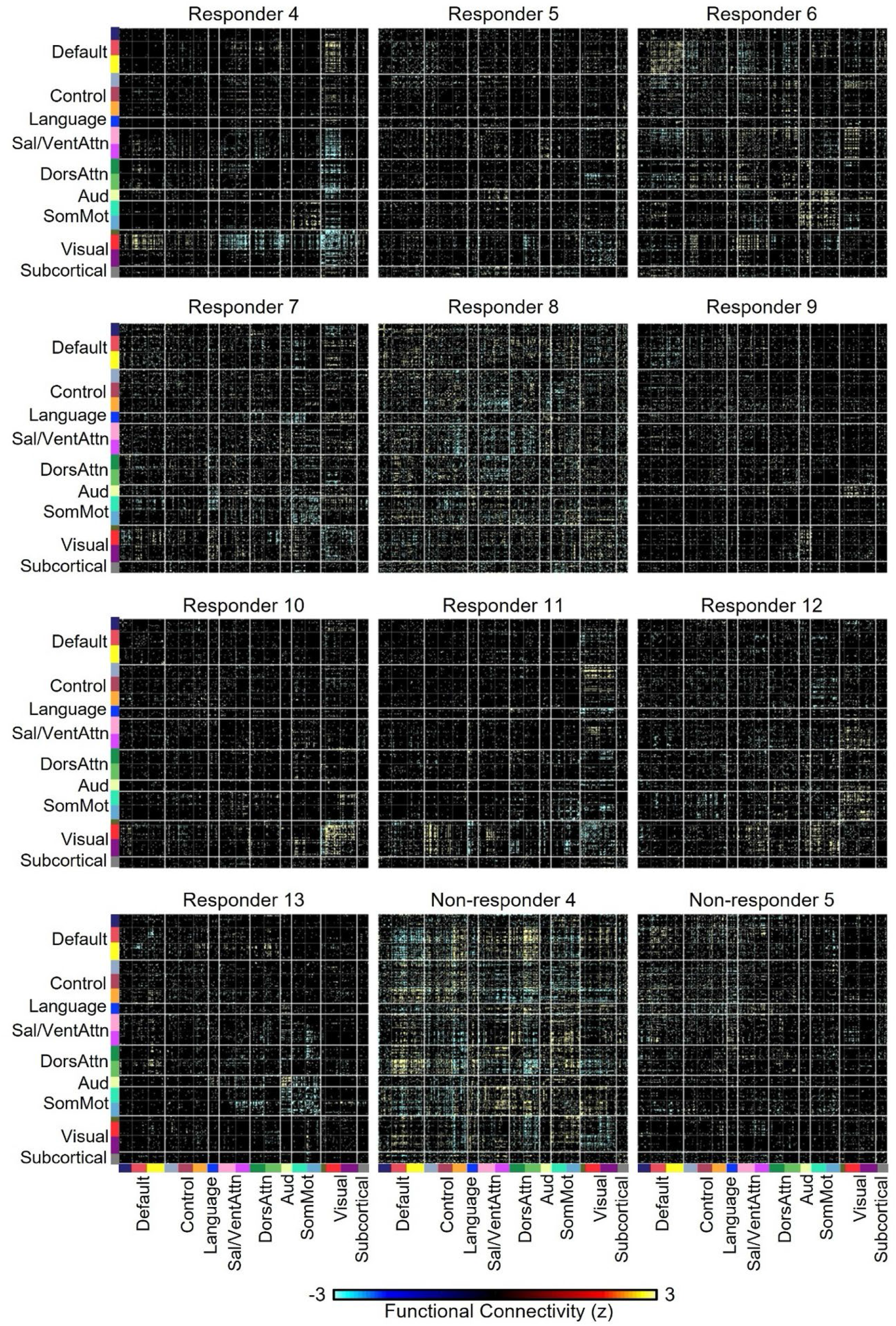
Thresholded functional connectivity (FC) change matrices for the remaining participants, with edges surviving false discovery rate (FDR) correction at q < 0.05 highlighted.

**Figure S8.**
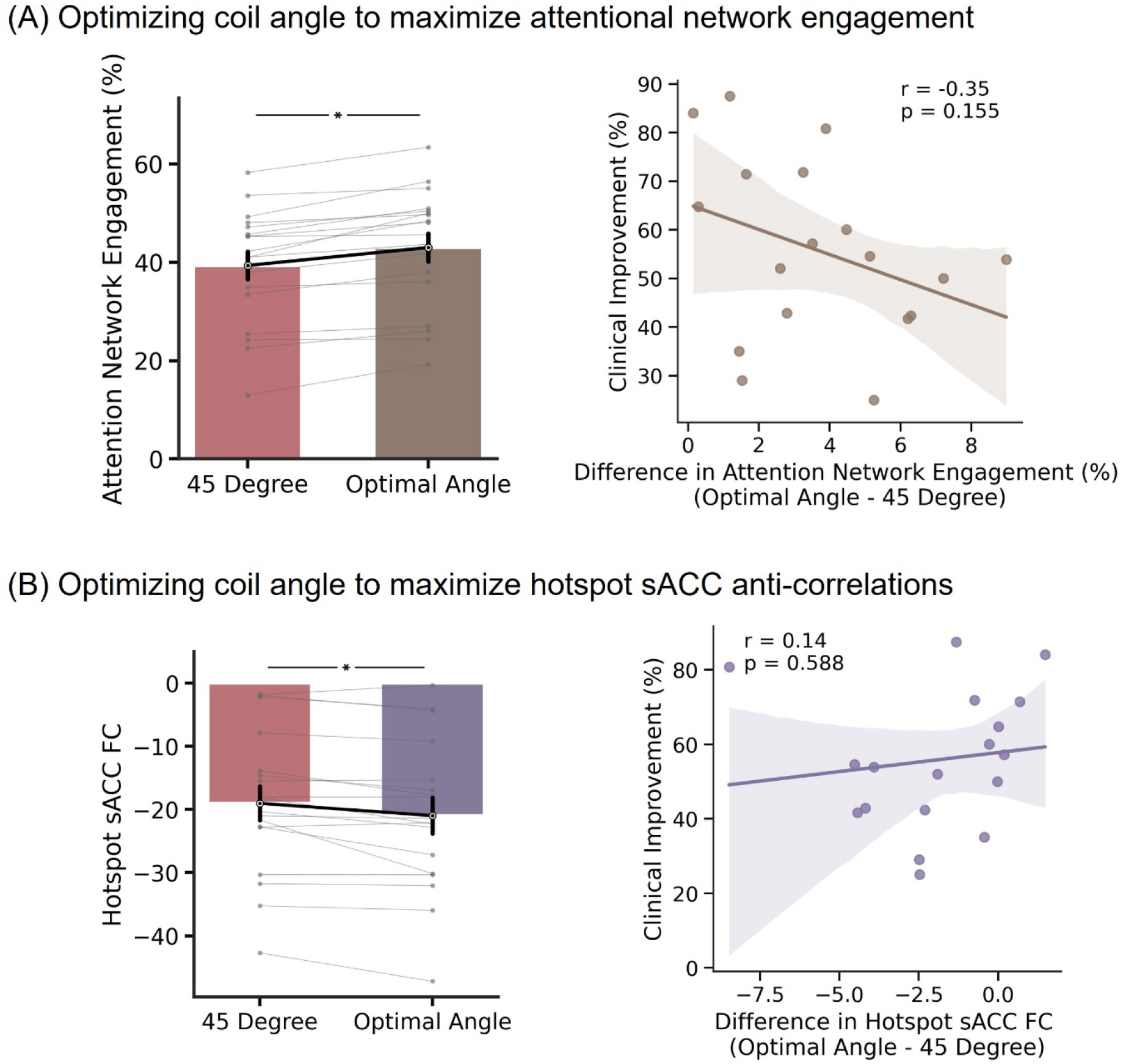
Post hoc exploratory analysis of the impact of coil orientation on clinical efficacy. (A) Optimizing coil angle to maximize attentional network engagement. The left panel shows the attention network engagement for the actual coil angle (45 degrees) and the optimal coil angle. The right panel shows the correlation of percentage MADRS improvement with the differences in on-target attentional network engagement between optimal and actual coil angles. (B) Optimizing coil angle to maximize hotspot sACC anti-correlations. The left panel shows the hotspot sACC FC for the actual coil angle (45 degrees) and the optimal coil angle. The right panel shows the correlation of percentage MADRS improvement with the differences in hotspot sACC FC between optimal and actual coil angles. Both correlations were directionally consistent with a possible clinical benefit of optimizing coil orientation, but neither reached statistical significance in this small sample.

**Figure S9.**
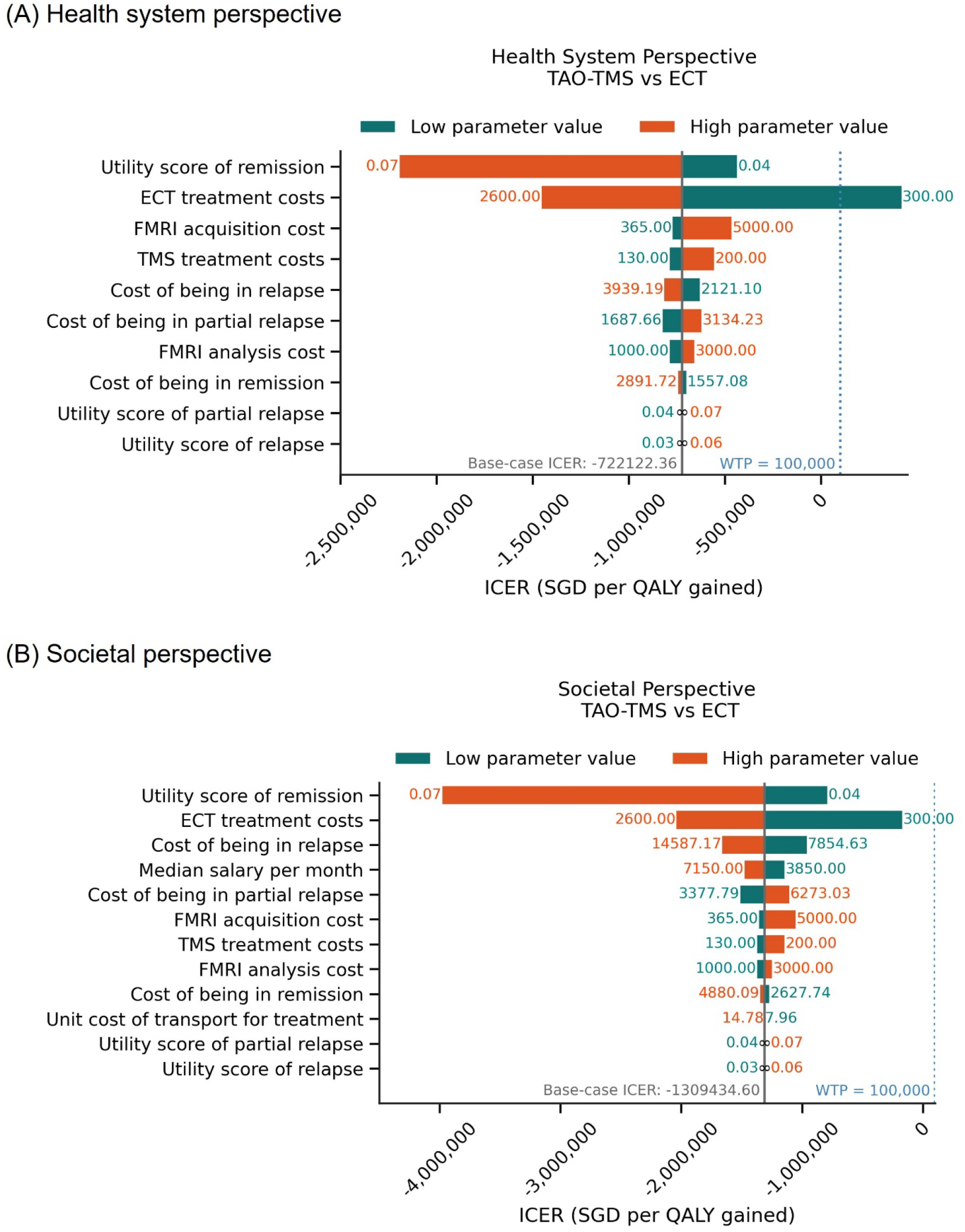
Tornado diagrams illustrating the results of one-way sensitivity analyses. The impact of individual input parameters on cost-effectiveness from (A) the health system perspective and (B) the societal perspective. Longer bars indicate greater influence of the parameter on the incremental cost-effectiveness ratio (ICER).

**Figure S10.**
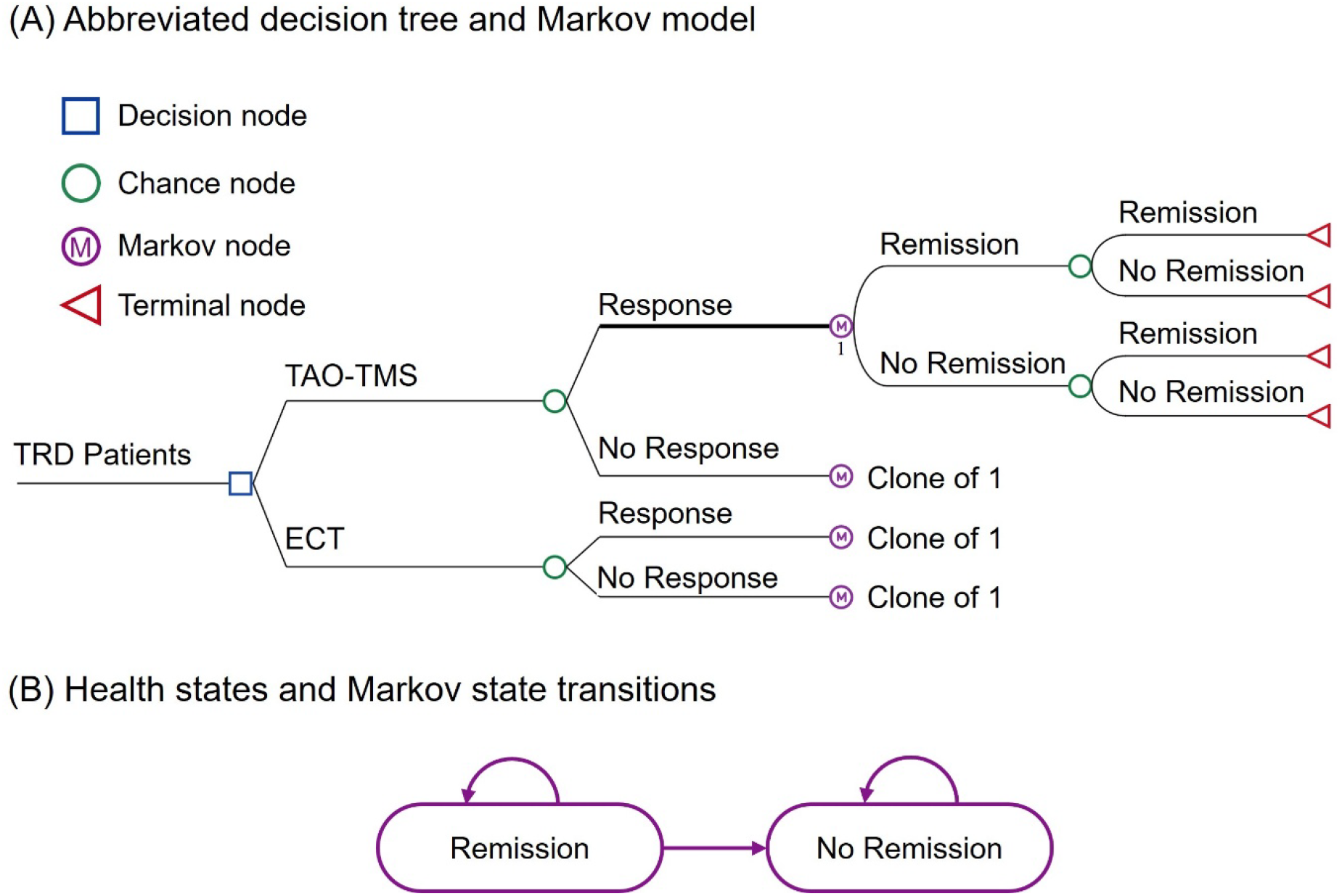
Decision analytical model comparing TAO-TMS and ECT for scenario analysis with two treatment-response categories and two Markov health states. (A) Abbreviated decision tree and Markov model representing the clinical pathways for patients receiving TAO-TMS or ECT. Initial treatment decisions from the decision tree fed into the Markov model for long-term simulation of outcomes and costs. (B) Health states and Markov state transitions for two health states. Arrows indicate possible transitions between health states at each cycle, reflecting treatment remission or no remission for each intervention. No remission was modeled as an absorbing state, with no transition from no remission to remission.

